# Machine Learning-Based Reconstruction of 2D MRI for Quantitative Morphometry in Epilepsy

**DOI:** 10.1101/2024.06.24.24309298

**Authors:** Corey Ratcliffe, Peter N. Taylor, Christophe de Bézenac, Kumar Das, Shubhabrata Biswas, Anthony Marson, Simon S. Keller

## Abstract

**Introduction:** Structural neuroimaging analyses require ‘research quality’ images, acquired with costly MRI acquisitions. Isotropic (3D-T1) images are desirable for quantitative analyses, however a routine compromise in the clinical setting is to acquire anisotropic (2D-T1) analogues for qualitative visual inspection. ML (Machine learning-based) software have shown promise in addressing some of the limitations of 2D-T1 scans in research applications, yet their efficacy in quantitative research is generally poorly understood. Pathology-related abnormalities of the subcortical structures have previously been identified in idiopathic generalised epilepsy (IGE), which have been overlooked based on visual inspection, through the use of quantitative morphometric analyses. As such, IGE biomarkers present a suitable model in which to evaluate the applicability of image preprocessing methods. This study therefore explores subcortical structural biomarkers of IGE, first in our ‘silver standard’ 3D-T1 scans, then in 2D-T1 scans that were either untransformed, resampled using a classical interpolation approach, or synthesised with a resolution and contrast agnostic ML model (the latter of which is compared to a separate model).

**Methods:** 2D-T1 and 3D-T1 MRI scans were acquired during the same scanning session for 33 individuals with drug-responsive IGE (age *mean* 32.16 ± *SD* = 14.20, male *n* = 14) and 42 individuals with drug-resistant IGE (31.76 ± 11.12, 17), all diagnosed at the Walton Centre NHS Foundation Trust Liverpool, alongside 39 age- and sex-matched healthy controls (32.32 ± 8.65, 16). The untransformed 2D-T1 scans were resampled into isotropic images using NiBabel (res-T1), and preprocessed into synthetic isotropic images using SynthSR (syn-T1). For the 3D-T1, 2D-T1, res-T1, and syn-T1 images, the recon-all command from FreeSurfer 8.0.0 was used to create parcellations of 174 anatomical regions (equivalent to the 174 regional parcellations provided as part of the DL+DiReCT pipeline), defined by the aseg and Destrieux atlases, and FSL `run_first_all` was used to segment subcortical surface shapes. The new ML FreeSurfer pipeline, `recon-all-clinical`, was also tested in the 2D-T1, 3D-T1, and res-T1 images. As a model comparison for SynthSR, the DL+DiReCT pipeline was used to provide segmentations of the 2D-T1 and res-T1 images, including estimates of regional volume and thickness. Spatial overlap and intraclass correlations between the morphometrics of the eight resulting parcellations were first determined, then subcortical surface shape abnormalities associated with IGE were identified by comparing the FSL `run_first_all` outputs of patients with controls.

**Results:** When standardised to the metrics derived from the 3D-T1 scans, cortical volume and thickness estimates trended lower for the 2D-T1, res-T1, syn-T1, and DL+DiReCT outputs, whereas subcortical volume estimates were more coherent. Dice coefficients revealed an acceptable spatial similarity between the cortices of the 3D-T1 scans and the other images overall, and was higher in the subcortical structures. Intraclass correlation coefficients were consistently lowest when metrics were computed for model-derived inputs, and estimates of thickness were less similar to the ground truth than those of volume. For the people with epilepsy, the 3D-T1 scans showed significant surface deflations across various subcortical structures when compared to healthy controls. Analysis of the 2D-T1 scans enabled the reliable detection of a subset of subcortical abnormalities, whereas analyses of the res-T1 and syn-T1 images were more prone to false-positive results.

**Conclusions:** Resampling and ML image synthesis methods do not currently attenuate partial volume effects resulting from low through plane resolution in anisotropic MRI scans, instead quantitative analyses using 2D-T1 scans should be interpreted with caution, and researchers should consider the potential implications of preprocessing. The `recon-all-clinical` pipeline is promising, but requires further evaluation, especially when considered as an alternative to the classical pipeline.

**Key Points:**

- Surface deviations indicative of regional atrophy and hypertrophy were identified in people with idiopathic generalised epilepsy.
- Partial volume effects are likely to attenuate subtle morphometric abnormalities, increasing the likelihood of erroneous inference.
- Priors in synthetic image creation models may render them insensitive to subtle biomarkers.
- Resampling and machine-learning based image synthesis are not currently replacements for research quality acquisitions in quantitative MRI research.
- The results of studies using synthetic images should be interpreted in a separate context to those using untransformed data.

## 1 Introduction

### 1.1 Clinical data

In the clinical evaluation pathway of non-specialist centres (i.e. general hospitals in the UK), research quality imaging data is not always feasible or accessible. Because of this, there is a growing amount of legacy data on PACS systems that remains relatively under-explored. (Papanicolas et al., 2018) There is also a growing interest in sustainability within science, which extends to minimising research waste, and the utility of repurposed clinical data for quantitative neuroimaging analyses is axiomatic. (Samuel et al., 2022) Many of the benefits associated with open-source datasets also apply to methods that increase the usability of retrospective clinical data, such as reducing research costs and increasing accessibility - repurposed clinical MRI data can be anonymised, distributed, and used by researchers who may not otherwise have access to it. (Laird, 2021; Madan, 2022) Access to large amounts of neuroimaging data is especially valuable in the advent of big data approaches. (Van Horn & Toga, 2014) Alongside (and partly due to) the popularisation of big data research, Machine-learning based (ML) technologies have also seen adoption within and beyond neuroimaging, emphasised by the recent release of FreeSurfer’s `recon-all-clinical` (RC) pipeline. (Billot, Greve, et al., 2023; Billot, Magdamo, et al., 2023; Gopinath et al., 2023; Iglesias et al., 2023; LeCun et al., 2015)

With the proliferation of capable hardware and software, MRI analyses have become a common method for exploring structural correlates of various pathologies, providing insights into both the prognostication and characterisation of neurological disease. There is, however, a trade-off between spatial resolution and ease of acquisition, which can influence quantitative analysis. (Brownhill et al., 2021; Despotović et al., 2015; Vidal-Jordana et al., 2017) It is a recognised limitation of MRI research that analyses are dependent on input data quality, and deviations from the research ideal (i.e. near-isotropic 1mm MP-RAGE, a sequence that analysis software is often designed to favour) are detrimental to the reliability of the results - indeed, it is common for processing software to specify that results obtained from anisotropic data may be inaccurate, as is the case for both FreeSurfer’s `recon-all` (RA) pipeline, and DL+DiReCT. (Edelman et al., 2009; Fischl, 2012; Mugler III & Brookeman, 1990; Rebsamen et al., 2022)

Partial Volume Effects (PVE), error introduced by the difficulty in deconvolving multiple tissue types from a single voxel intensity, which can lead to misclassification of tissue, are more pronounced when voxels are larger. (Firbank et al., 1999) Whilst qualitative inspection by experts is less susceptible to systematic bias resulting from acquisition parameters than quantitative analyses are, anisotropic voxels are differentially susceptible to PVE in different orientations, leading to a systematic bias when estimating, for example, the the volume of the cortical surface. (Lie et al., 2022; Rebsamen et al., 2023) In the subcortical structures, a prior study demonstrated that the consistency of volume estimations between isotropic (3D-T1) and anisotropic (2D-T1) images decreased proportional to the degree of anisotropy. (Brownhill et al., 2021)

Low resolution scans are therefore not ideal for brain morphometry and deviations of basic imaging parameters (i.e. voxel size) can lead to systematic bias. (Haller et al., 2016; May & Gaser, 2006; Tor-Díez et al., 2019) Whilst the in plane resolution of the images can be increased without much consequence to the patient/clinician, acquiring data in 3D can significantly increase scan times, which increases cost and patient discomfort. Long scans are undesirable, so there is an argument for increasing the step size (and thus improving patient experience/reducing artifact susceptibility) through plane, and creating anisotropic voxels in an attempt to facilitate qualitative visual analysis/increase diagnostic yield of the scan without overextending resources. (Mayberg et al., 2022)

### 1.2 Processing clinical data

Several methods for coercing clinical imaging data into a more homogenous format (i.e. isotropic) have been proposed, tested, and in some cases even widely adopted. There has been, however, no comparative systematic evaluation of these methods with reference to same session intrasubject 3D-T1 ‘ideal’ scans. This paper will therefore evaluate the ability of resampling and various ML models to improve upon the coherence of outputs from the quantitative analyses of 2D-T1 scans with those from analogous 3D-T1 scans.

The first and simplest method, resampling, is a context-naïve mathematical operation used to split or combine *k* pre-existing voxels into *j* voxels of arbitrary dimensions, using interpolation algorithms to estimate values which satisfy the (unknown) midpoints between two known voxel values. (Li et al., 2004) The conform command in the Nibabel software suite is a popular method of accomplishing this. (Brett et al., 2023) Resampling can be used to create the impression that an image is isotropic by splitting voxels, however, resampling cannot resolve the systematically biased PVE introduced by an anisotropic acquisition. Nonetheless, it is a common neuroimaging analysis technique that has a myriad of uses, including preparing anisotropic scans for morphometry with the DL+DiReCT tool, one of the ML models used in this paper.

Briefly, machine learning is a branch of artificial intelligence wherein algorithms are developed from a subset of data, and extrapolated to represent a broader range of input/outcome operations. (Choi et al., 2020; Currie et al., 2019) When an ML model contains multiple hidden computational layers, it is considered a deep learning model. Developed for use with contrast-enhanced clinical MRI scans, DL+DiReCT uses a proprietary deep learning model, DeepSCAN, to segment and parcellate tissue classes in images with systematic bias, before estimating cortical thickness using the `antsCorticalThickness.sh` tool. (Brett et al., 2023; Das et al., 2009; McKinley et al., 2021; Tustison et al., 2013) Through the implementation of a U-net CNN (Convolutional Neural Network) trained on parcellations from a large training data set, DeepSCAN uses generalised paired information on the cortical structure (i.e. context priors) to robustly segment tissue types (grey matter, white matter), ameliorating the uncertainty introduced by white matter lesions and the application of a contrast-enhancing agent. (Rebsamen et al., 2020, 2022) DL+DiReCT offers three distinct pre-trained models for use in the segmentation algorithm, one of which (v0) is trained on manually segmented scans from healthy volunteers and people with various neurological conditions, whilst the other two are enriched with data from a cohort of multiple sclerosis patients, offering two distinct atlas choices (v6, Desikan-Killiany; v7, Destrieux).

SynthSR is an input-agnostic ML joint super-resolution and contrast synthesis model, which uses a 3D U-net CNN to create 1mm isotropic MP-RAGE versions of input images. More specifically, a model that is independently trained on paired images estimates an isotropic analogue of the (unpaired) input using context priors (from the paired training images) to supplement the information in the input, and then a model trained on source and target modality pairs (augmented with a further Generative Adversarial Network model to supplement ‘real scan’ pairs with synthetic ones, thereby enhancing the training data) re-estimates the tissue response intensities that would be expected in an MP-RAGE scan. (Iglesias et al., 2023) This approach is novel for its ability to compute super-resolution and contrast synthesis of an image without paired input data (i.e. explicitly coupled source and target images), its ability to combine data from multiple heterogenous scans, and its ability to synthesise a reference contrast (MP-RAGE). (Billot, Greve, et al., 2023; J. Du et al., 2020; Huang et al., 2017; Zhao et al., 2021)

### 1.3 Structural biomarkers in idiopathic generalised epilepsy

The current understanding of epilepsy—one of the most common neurological diseases globally—as a network disorder has been heavily influenced by the results of structural MRI studies based on prospectively acquired, research quality data. (Beghi, 2020) Patterns of volumetric, morphometric, and network abnormalities have been associated with common, syndrome-specific, and prognostic phenotypes of epilepsy. (Bonilha & Keller, 2015; Hatton et al., 2020; Keller et al., 2012, 2015; Larivière et al., 2020; Whelan et al., 2018) People with epilepsy routinely present with marked, yet subtle, patterns of subcortical atrophy that may not appear visually abnormal on clinical MRI, even at the point of diagnosis. (Leek et al., 2021)

Atrophy of the subcortical structures has been repeatedly observed in cohorts of individuals with Idiopathic Generalised Epilepsy (IGE), potentially related to syndrome-specific pathomechanisms; abnormalities of the hippocampi, putamen, and thalami are of particular relevance to epileptogenesis, constituting biomarkers of IGE. (Caciagli et al., 2019; H. Du et al., 2011; Keller et al., 2011) Furthermore, the relationship between structural abnormalities and the phenotype of IGE extends beyond seizure activity to encompass cognitive impairment, indicating system-wide network reorganisation. (Ratcliffe et al., 2020) The reliability of these structural correlates makes them an appropriate model for testing the veracity of image processing and manipulation.

### 1.4 Objectives

In the present study we systematically evaluate methods for optimising quantitative structural analysis when using clinical imaging data, i.e. which analogous (natively anisotropic) image provides the measurements and metrics with the closest coherence to the ‘fit for purpose’ 3D-T1 scan: the untransformed 2D-T1 image, the 2D-T1 image resampled with NiBabel, the ML super-resolution MP-RAGE image generated with SynthSR (an input-agnostic model), or 2D-T1 image segmented and parcellated with DL+DiReCT (using a model robust to artifacts present in contrast-enhanced multiple sclerosis data).

We quantify coherence through comparison of the analogue-derived volumetry and morphometry with that from the silver-standard 3D-T1 scan. With this data, we first determine within-subject variation, and then explore IGE-related subcortical surface shape abnormalities using our analogous input images. Our study follows numerous investigations into the efficacy of model-based image synthesis, alongside other methods with more specific utility—such as super-resolution of isotropic images, or in neonates. (Delannoy et al., 2020; Fiscone et al., 2024; Iglesias et al., 2021; Rebsamen et al., 2022; Rusak et al., 2022; Tian et al., 2021) This is, however, the first independent investigation into, and comparison of, the relative efficacy of several common and generalisable artifact compensation methods. To help neuroimagers make a more informed choice, we demonstrate the relative caveats and benefits of several potential structural preprocessing methods.

## 2 Materials and Methods

### 2.1 Participants and scanning protocols

This study was conducted using participants and data that have been previously described in truncated form. (Brownhill et al., 2021; McKavanagh et al., 2021, 2023; Pegg et al., 2021) All participants provided informed written consent, and data collection was approved by the local ethics committee (UK Research Ethical Committee ref. 14/NW/0332). 31 (33 before exclusions) individuals with drug-responsive IGE (age *mean* 32.16 ± *SD* = 14.20, male *n* = 14; pwIGE) and 42 individuals with drug-resistant IGE (31.76 ± 11.12, 17; pwDRE), all diagnosed at the Walton Centre NHS Foundation Trust Liverpool, were recruited through referral from the examining clinician. 39 age- and sex-matched Healthy Controls (32.32 ± 8.65, 16; HC) were also recruited from around the Liverpool/Aintree area after being approached by researchers.

For all people with Idiopathic Generalised Epilepsy (pwE) in our cohort, diagnoses of IGE were made by an epileptologist based on semiology, EEG characteristics, and clinical history, and are consistent with current International League Against Epilepsy guidelines. (Scheffer et al., 2017) Drug-resistance was determined based on whether the patient had been seizure free with the use of Anti-seizure Medication (ASM) for the six months preceding their last follow-up or not. Pertinent demographic information was recorded at the time of recruitment. Full sample characteristics are presented in Appendix 1.

Scanning took place in a GE Discovery MR750 3.0T MRI scanner at the Walton Centre NHS Foundation Trust between 2014 and 2016. For each participant, two scans were acquired over a single session: a 3D T1-FSPGR PURE (Fast Spoiled Gradient Echo with Phased Array Uniformity Enhancement signal inhomogeneity correction) with TR = 8.2ms, TI = 450ms, TE = 3.22ms, flip angle = 12°, voxel size = 0.9 × 0.9 × 1.0mm, acquisition matrix = 256 × 256 × 135, and FOV = 240 × 240 × 135mm; and a 2D T1-FLAIR (Fluid Attenuated Inversion Recovery) with TI = 920ms, TE = 9.94ms, flip angle = 111°, voxel size = 0.4 × 3.0 × 0.4mm, acquisition matrix = 512 × 52 × 396, and FOV = 220 × 156 × 170mm. See Figure 1 for a comparison of anisotropic and isotropic image acquisitions.

**Figure 1.**
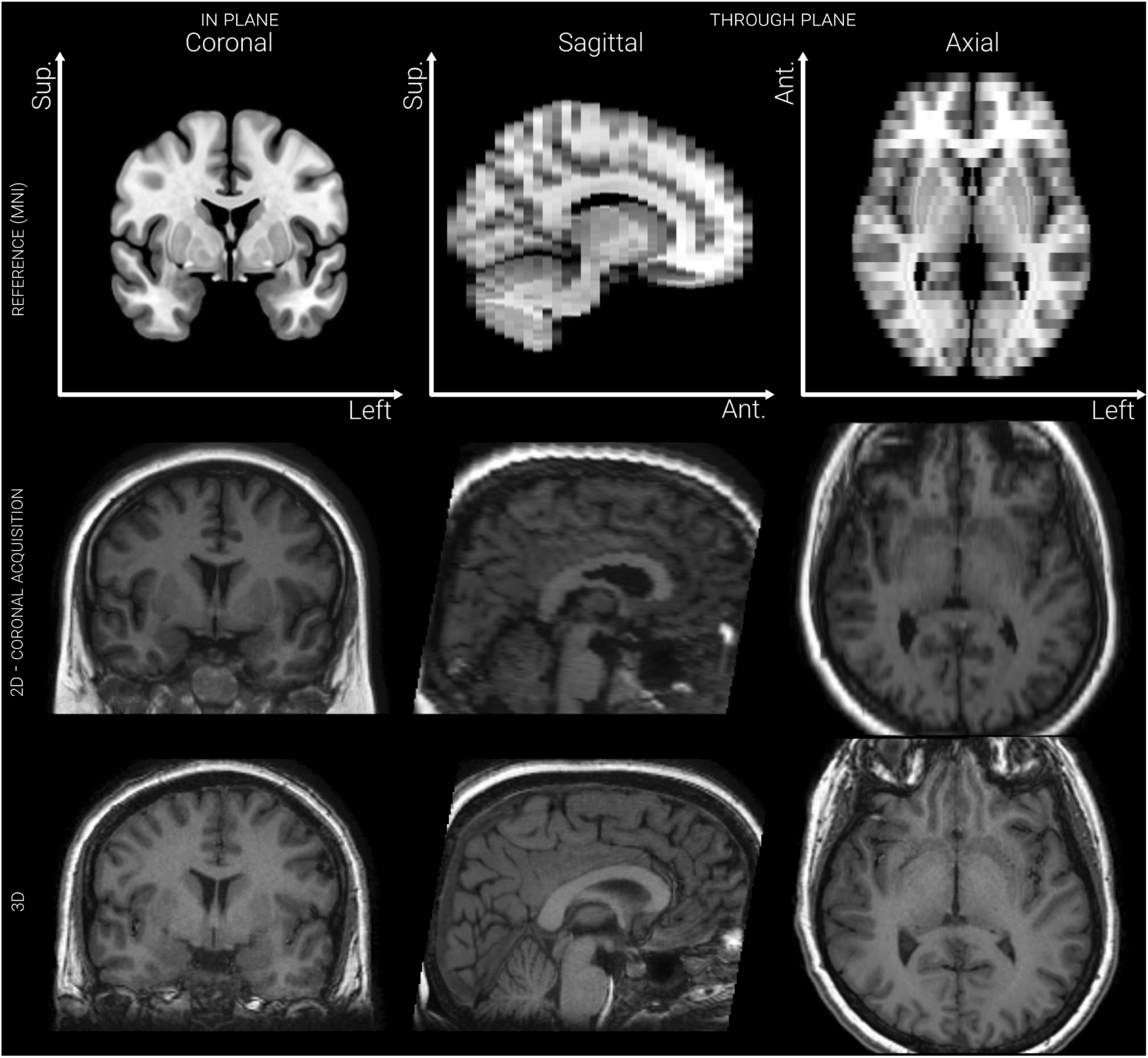
Slices from 2D-T1 and 3D-T1 MRI scans of the same participant, annotated with the planes defined by acquisition. Slices from an artificially anisotropic version of the MNI-space (Montreal Neurological Institute) 152 subject average brain MRI are also presented, to illustrate the effect of anisotropy.

### 2.2 Image synthesis and resampling

The code used for image synthesis, processing, and analysis is provided on GitHub. As shown in Figure 2a., the 2D-T1 scans were, in parallel, synthesised with SynthSR, or preprocessed using DL+DiReCT. For each 2D-T1 scan, the standalone implementation of SynthSR was run to predict a best-guess estimate of a isotropic MP-RAGE image (voxel size 1 × 1 × 1mm, matrix = 221 × 157 × 171), using a model based on the included training dataset. (Billot, Greve, et al., 2023; Iglesias et al., 2021)

**Figure 2.**
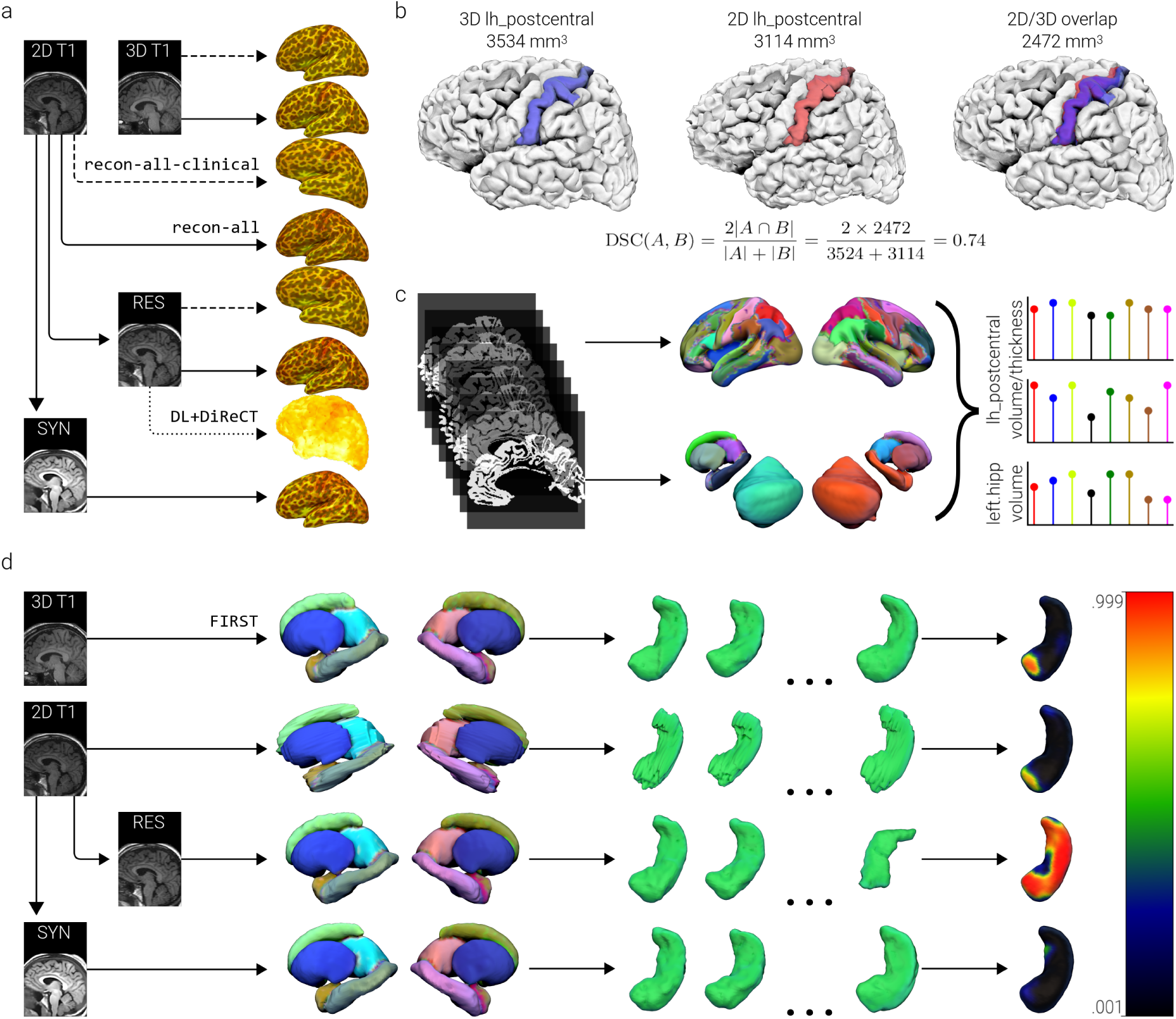
A visual schematic of the two image processing arms of the study: the FreeSurfer surface-based morphometry pipeline (a, b, c), and the FSL subcortical mesh and volumetry pipeline (d). a. The isotropic (3D-T1) images were processed with both `recon-all` and `recon-all-clinical` from the FreeSurfer software package, as were the raw (2D-T1) anisotropic images and the resampled (res-T1) images. The resampled images were also parcellated using the DL+DiReCT pipeline, and images generated using SynthSR (syn-T1) were parcellated with recon-all only. b. On a per-subject basis, spatial overlap was computed between the 3D-T1 scan parcellations and the seven analogue images, for each of the 174 regions in the truncated Destrieux atlas. To account for error in orientation between the scans, which may have been exacerbated during processing, the parcellations were registered to their isotropic analogue with a rigid-body alignment. DSC is sensitive to slight differences in location, as well as in shape. c. Volume and (for the cortical regions) thickness measurements were extracted from the parcellations and standardised based on the distribution of estimates for that region/metric pair in the isotropic images. d. The `run_first_all` pipeline was applied to the 3D-T1, 2D-T1, res-T1, and syn-T1 images, providing volumes and surface meshes for 14 subcortical structures (the brainstem was omitted). Non-parametric permutation testing was then used to explore surface mesh deformation (i.e. deflation and inflation) between our different cohorts, which could then be contrasted across image types. Note that one of the hippocampal meshes for the resampled data appears to be oriented incorrectly. Both scans were taken from the same subject, during the same session. DSC, Dice Similarity Coefficient

The resampled 2D image (voxel size 1 × 1 × 1mm, matrix = 256 × 256 × 256), which had been skull-stripped with HD-BET and resampled with nibabel.processing.conform as part of the DL+DiReCT pipeline, was extracted for parallel processing, as well as used as the input for the DeepSCAN (model v7) segmentation and parcellation step. (Brett et al., 2023; Isensee et al., 2019; McKinley et al., 2021) After ML segmentation, antsCorticalThickness was automatically applied to estimate cortical thickness and volume measurements for regions based on a modified version of the Destrieux atlas provided in FreeSurfer 5.3. (Das et al., 2009; Destrieux et al., 2010; Rebsamen et al., 2020, 2022; Tustison et al., 2013) After this stage, we had four structural images per participant, as well as DL+DiReCT metrics:

- 3D-T1: T1-FSPGR-PURE, acquired with a near-isotropic resolution
- 2D-T1: (untransformed) T1-FLAIR, acquired with an anisotropic resolution
- res-T1: Resampled (mathematically interpolated) and skull-stripped 2D-T1
- syn-T1: SynthSR derived pseudo-isotropic T1
- dl-T1: The quantitative outputs of the DL+DiReCT pipeline

### 2.3 Image preprocessing

To concisely reflect structural neuroimaging clinical research, our comparisons were limited to the clinically interpretable volume and thickness measurements of the cortical parcellations, and volumes/surfaces of the subcortical regions. We present a schematic overview of the image processing pipelines in Figure 2. Assessment of volumetric and morphometric measurements began with the reconstruction of tissue surfaces across all four image types and 112 participants using the recon-all (RA) pipeline in FreeSurfer 8.0.0. (Fischl, 2012) The newly-released `recon-all-clinical` (RC) pipeline was also applied to the 3D-T1, 2D-T1, and res-T1 images. The resulting parcellations were subsequently registered to the subject-specific 3D-T1 parcellation using `flirt` with 6 degrees-of-freedom (a rigid-body registration, allowing for rotation and translation) and nearest-neighbour interpolation.

From the DL+DiReCT pipeline, regional volume and thickness measurements for ML segmentations based on the res-T1 images were also available. (Das et al., 2009; McKinley et al., 2021) To assure consistent comparisons between processing arms, parcellations redefined by the more recent releases of FreeSurfer were edited to match those used by DL+DiReCT (i.e. subdivisions in the cerebellum and corpus callosum were summed, and seven regions with no direct regional basis were omitted). The final parcellation maps contained 148 cortical regions and 26 subcortical segmentations.

To evaluate the ability of resampling and synthesis to compensate for resolution-based information loss when distinguishing between groups, subcortical surface volumes and shapes were compared between HC and pwE (then between HC, pwDRE, and pwIGE, see Appendix 2 and 3) first in the 3D-T1 scans, and then in the analogues (i.e. the 2D-T1, res-T1, and syn-T1 images). For subcortical volumetry and shape analysis, all four structural images across all 112 participants were first bias-corrected with the `N4BiasFieldCorrection` algorithm, brain extracted with the BET tool, linearly registered to the brainextracted MNI template with FLIRT, and then segmented and modelled with the `run_first_all` pipeline (from the Advanced Normalization Tools [ANTs] library and FMRIB Software Library [FSL], respectively). (Jenkinson & Smith, 2001; Jenkinson et al., 2002, 2012; S. Smith, 2002; Tustison et al., 2010)

### 2.4 Statistical analyses

To quantify the similarity of the image analogues with the ‘silver-standard’ ground truth represented by the 3D-T1 scans (gold standard, in this case, would require direct *ex-vivo* measurement), Dice Similarity Coefficients (DSC) across all parcellations were computed using `fslmaths` to first create ‘overlap’ images; a demonstration of how DSC can be used to assess regional overlap is provided in Figure 2b. (Jenkinson et al., 2012; Yao et al., 2020) Further comparisons of cortical metrics were carried out in R, the details of which (including packages, and visualisation tools) are presented in Appendix 4. (R Core Team, 2023; RStudio Team, 2020) Descriptive assessment of the deviation between the quantitative metrics derived from the analogue images were carried out after standardisation (i.e. z-scoring) of the cortical volumes, cortical thicknesses, and subcortical volumes to distributions defined by the respective 3D-T1 image metric (i.e. the left hippocampus volumes in all image types was z-scored to the distribution of left hippocampal volumes derived from the 3D-T1 scans; see Figure 2c.). Similarity between the metric datasets were measured using Intraclass Correlation Coefficients (ICC).

Subcortical volumes were compared between pwE and HC in R, with general linear models (GLMs), including age, sex, and Total Parenchymal Volume (TPV) as covariates. Following the recommended pipeline, subcortical surface shape comparisons between pwE and HC were also modelled via GLM (with the same covariates) using the FSL randomise permutation testing tool (*nperm* = 5000; all covariates standardised, according to FSL recommendations), and threshold free cluster enhancement for multiple comparisons. (S. M. Smith & Nichols, 2009) The same methods were also used to examine differences between sub-groups: HC, pwDRE, pwIGE (which are presented in Appendix 2 and Appendix 3). As any clinical interpretations were secondary to evaluating the methods involved, multiple comparison corrections were not applied between inferential tests (i.e. between subcortical structures or image types).

## 3 Results

### 3.1 Descriptive volumetric and morphometric comparisons

Standardised grey matter volume and thickness measurements derived from the analogue images were consistently lower than those recorded in the 3D-T1 scans, as shown in table 1, whereas deviations in the estimates of the subcortical volumetry were typically larger than the reference, and more subtle. Metrics computed using the untransformed 2D-T1 scans deviated the least from our ground truth (performing comparably to the 3D-T1 scans processed with `recon-all-clinical`, whereas the dl-T1 and syn-T1 metrics tended to deviate the most. Figure 3 presents scatterplots showing the variability of measurements from the analogue image relative to the same data from the 3D-T1 scans, highlighting the abnormally large variability of the cortical dl-T1 thickness estimates, and the lack of any systematic bias in the estimation of metrics related to region size in the analogue images. For wholebrain measures, see Appendix 3.

**Figure 3.**
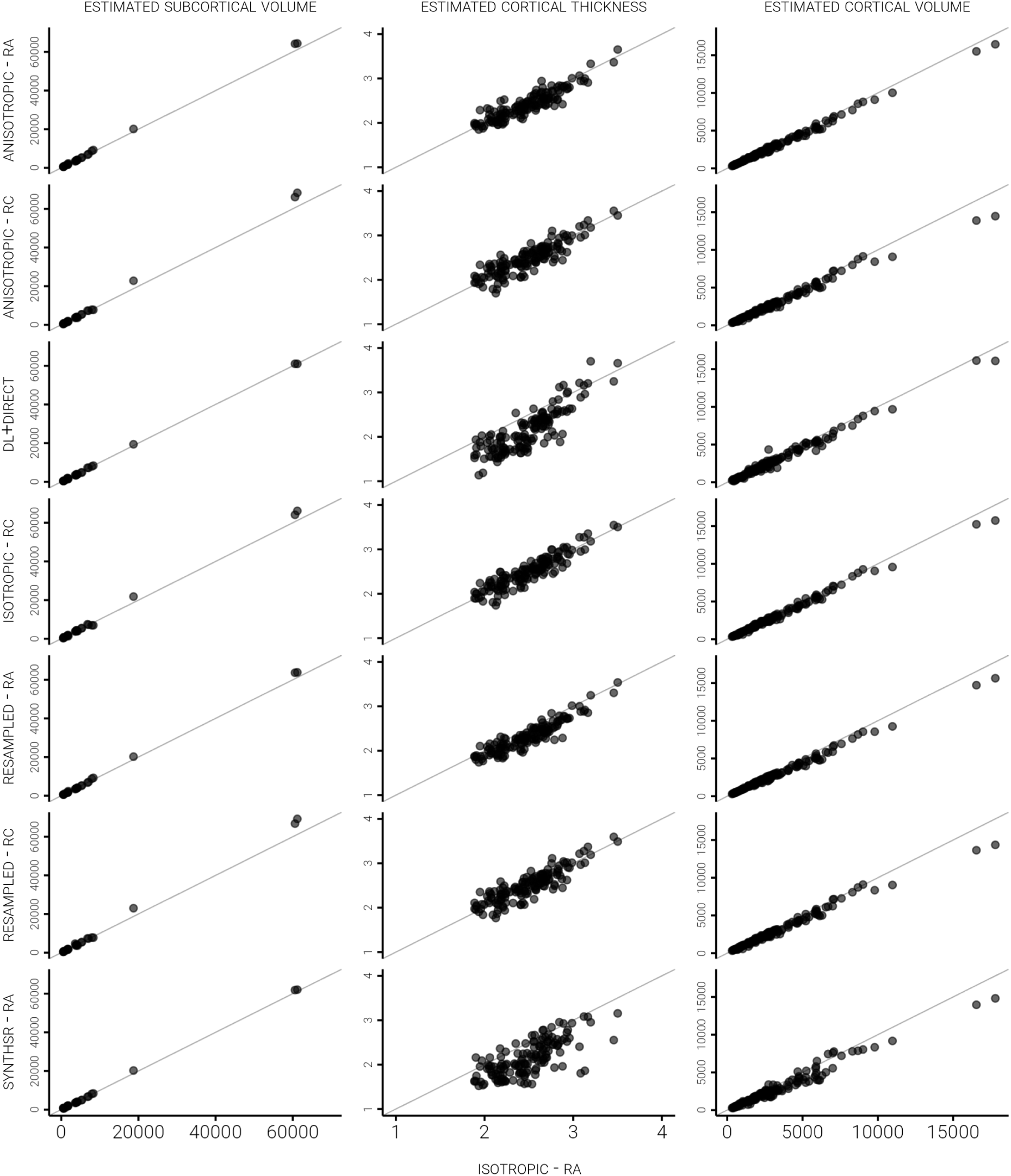
Scatterplots representing the relationships between FreeSurfer-extracted volume and thickness measurements of 148 parcellated cortical regions and 26 subcortical regions in the 3D-T1 scans (x-axes) and the seven analogue images (y-axes). A reference line for perfect agreement (i.e. y = x) is also plotted, demonstrating that whilst agreement is generally good between the image types, SynthSR and DL+DiReCT see greater deviation from the expected measures.

**Table 1.**
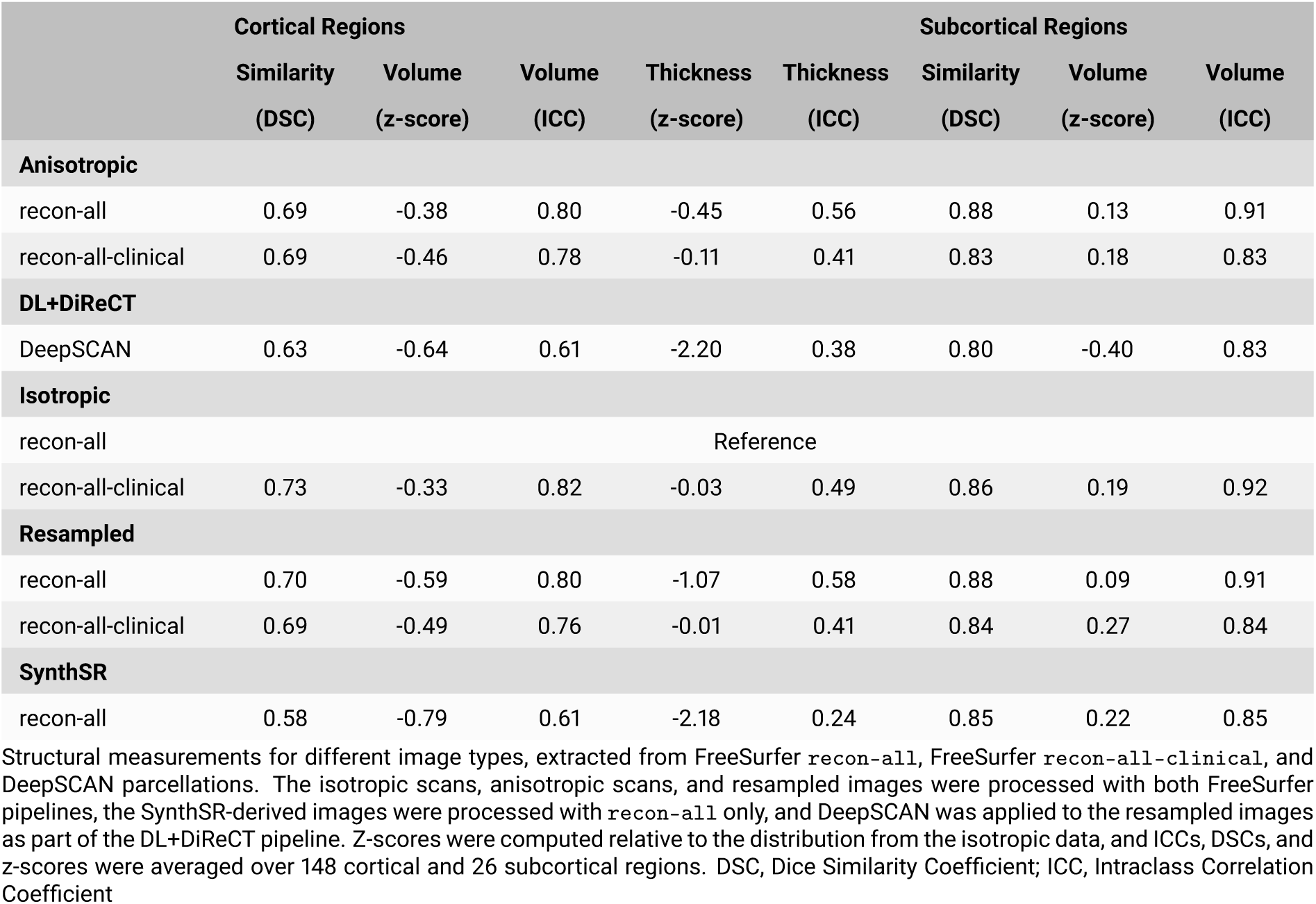
Regional descriptive summary stats.

### 3.2 Regionwise spatial correspondence

DSC were computed to quantify the spatial overlap between the 174 regions/segmentations of the 3D-T1 scans and the analogue images (Figure 4). Overlap for the 148 grey matter regions was middling-to-large on average (2D-T1 RA = 0.69, 2D-T1 RC = 0.69, dl-T1 = 0.63, 3D-T1 RC = 0.73, res-T1 RA = 0.70, res-T1 RC = 0.69, syn-T1 RA = 0.58; see table 1), and slightly larger for the 26 subcortical segmentations (2D-T1 RA = 0.88, 2D-T1 RC = 0.83, dl-T1 = 0.80, 3D-T1 RC = 0.86, res-T1 RA = 0.88, res-T1 RC = 0.84, syn-T1 RA = 0.85). The parcellations defined in the 2D-T1 and res-T1 images were the most comparable to those of the 3D-T1, whereas the dl-T1 and syn-T1 parcellations were consistently the least. The use of FreeSurfer’s RC pipeline had either no effect, or a small negative effect on regional overlap.

**Figure 4.**
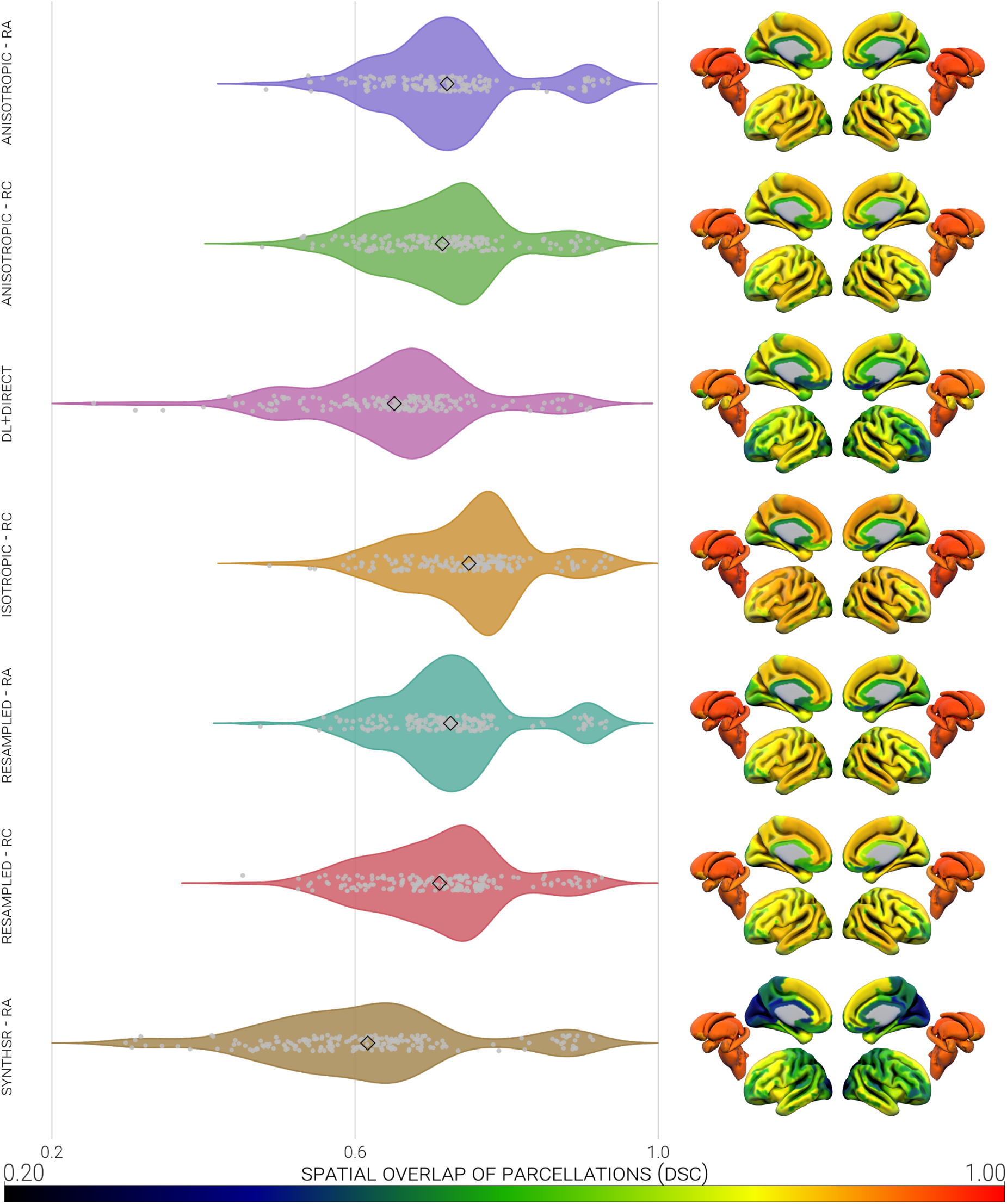
Heatmap and violin plot representations of the spatial overlap (presented on the scale of 0.2 to 1.0) of 174 parcellated regions created from 3D-T1 scans and analogues. Regional data were averaged across all participants in each image type, to leave one datum per region per image type. DSC, Dice Similarity Coefficient; RA, `recon-all`; RC, `recon-all-clinical`

### 3.3 Metric correspondence across image types

To quantify the similarity of the metrics extracted from each image type, ICC were computed between the 3D-T1 scan metrics and each of the analogue image metrics (FreeSurfer RA, RC, and DL+DiReCT included). For cortical volume (2D-T1 RA = -0.38, 2D-T1 RC = -0.46, dl-T1 = -0.64, 3D-T1 RC = -0.33, res-T1 RA = -0.59, res-T1 RC = -0.49, syn-T1 RA = -0.79; see table 1), cortical thickness (2D-T1 RA = -0.45, 2D-T1 RC = -0.11, dl-T1 = -2.20, 3D-T1 RC = -0.03, res-T1 RA = -1.07, res-T1 RC = -0.01, syn-T1 RA = -2.18), and subcortical volume (2D-T1 RA = 0.13, 2D-T1 RC = 0.18, dl-T1 = -0.40, 3D-T1 RC = 0.19, res-T1 RA = 0.09, res-T1 RC = 0.27, syn-T1 RA = 0.22), the 2D-T1 metrics had the highest correlation with those from the 3D-T1 scans, followed by those from the res-T1 images. The syn-T1 and dl-T1 values were the least similar to the 3D-T1 in all three included measures, appearing susceptible to outlier effects. The RC pipeline follows the trend of underestimating subcortical metrics, whilst ameliorating some of the variance in the thickness estimations. Volume measurements were less varied (relative to our ground truth) than thickness (Appendix 3).

### 3.4 Demographics

Two pwIGE were excluded prior to the study due to subsequent re-diagnosis, resulting in a sample of 42 pwDRE, 31 pwIGE and 39 HC. The groups did not significantly differ in age, sex, or TPV. Demographic and clinical characteristic summaries are presented in table 2, alongside the results of difference tests.

**Table 2.** Demographics.

|  | Healthy controls |  | People with drug-resistant IGE |  | People with drug-responsive IGE |  | *p* = |  |
| --- | --- | --- | --- | --- | --- | --- | --- | --- |
|  | *(n* = 39) |  | *(n* = 42) |  | *(n* = 31) |  |  |  |
| Age at scan in years mean (*SD*) | 32.38 | (8.65) | 31.76 | (11.12) | 32.16 | (14.20) | f-test: | .969 |
|  |  |  |  |  |  |  | HC/pwDRE: | .805 |
|  |  |  |  |  |  |  | HC/pwIGE: | .935 |
|  |  |  |  |  |  |  | pwIGE/pwDRE | .882 |
| TPV in mm<sup>3</sup> mean (*SD*) | 1583688 | (136239) | 1558160 | (176741) | 1594837 | (159805) | f-test: | .593 |
|  |  |  |  |  |  |  | HC/pwDRE: | .472 |
|  |  |  |  |  |  |  | HC/pwIGE: | .771 |
|  |  |  |  |  |  |  | pwIGE/pwDRE | .332 |
| Sex | 23/16 | (58.67%) | 25/17 | (59.52%) | 17/14 | (54.84%) | f-test: | .913 |
| F/M (*F*%) |  |  |  |  |  |  | HC/pwDRE: | .960 |
|  |  |  |  |  |  |  | HC/pwIGE: | .728 |
|  |  |  |  |  |  |  | pwIGE/pwDRE | .690 |

Sample demographics from the different groups within the cohort. For age at scan and TPV, differences in group variance were assessed with ANOVAs and uncorrected post-hoc pairwise t-tests. Differences in sex distributions were assessed with group and pairwise chi-square tests. TPV was calculated from the isotropic images, using FreeSurfer's `recon-all` command. IGE, Idiopathic Generalised Epilepsy; TPV, Total Parenchymal Volume

### 3.5 Subcortical surface morphometry

When HC and pwE were compared based on the 3D-T1 scans (derived by FSL `run_first_all`) there were uncorrected significant differences between pallidal volumes bilaterally (see table 3). The same differences were detected in the res-T1 data, whereas the 2D-T1 scans showed only differences in the volume of the right thalamus and the syn-T1 images indicated significant differences in the left accumbens, right hippocampus, and left thalamus. When subgroups were compared, there were no significant differences found in the isotropic data (see Appendix 2). Despite the absence of any differences in the ground truth data, several comparisons reached significance when calculated from the analogue-derived volumes: 14/56 from the res-T1 data, four from the syn-T1 data, and one from the 2D-T1 scans.

**Table 3.** Subcortical volume comparisons.

|  | Accumbens |  | Amygdala |  | Caudate |  | Hippocampus |  | Pallidum |  | Putamen |  | Thalamus |  |
| --- | --- | --- | --- | --- | --- | --- | --- | --- | --- | --- | --- | --- | --- | --- |
|  | L | R | L | R | L | R | L | R | L | R | L | R | L | R |
| **People with IGE vs. healthy controls** |  |  |  |  |  |  |  |  |  |  |  |  |  |  |
| Anisotropic | .188 | .063 | .685 | .298 | .543 | .284 | .876 | .247 | .097 | .327 | .225 | .330 | .134 | **.037** |
| Isotropic | .527 | .509 | .753 | .143 | .401 | .302 | .751 | .515 | **.047** | **.040** | .209 | .231 | .194 | .071 |
| Resampled | .131 | .295 | .944 | .767 | .408 | .614 | .691 | .767 | **.049** | **.040** | .698 | .234 | .492 | .422 |
| SynthSR | **.014** | .645 | .612 | .496 | .381 | .269 | .366 | **.012** | .410 | .990 | .092 | .257 | **.018** | .181 |

The uncorrected *p*-values from Kruskal-Wallis rank sum tests of subcortical volume differences (as measured using the FSL command, `run_first_all`) between people with IGE and healthy controls, observed across image types. **Bold** values and red cells highlight statistical significance at $\alpha = .050$ . Healthy Controls, *n* = 39; People with IGE, *n* = 73. IGE, Idiopathic Generalised Epilepsy

The subcortical shape differences commonly reported in IGE were observable in our sample (see table 4 and Figure 5) using the 3D-T1 data. Following permutation testing, clusters of significant inwards surface deflation (as a proxy of regional subcortical atrophy) were reported in the right caudate (*p corr* = .012), right pallidum (*p corr* = .025), left accumbens (*p corr* = .049), and bilateral thalamus (left *p corr* = .014, right *p corr* = .015) of our pwE cohort when compared to HC. Potential hypertrophy relative to HC (i.e. outwards surface inflation) was also identified in the right caudate (*p corr* = .018) of the pwE group.

**Figure 5.**
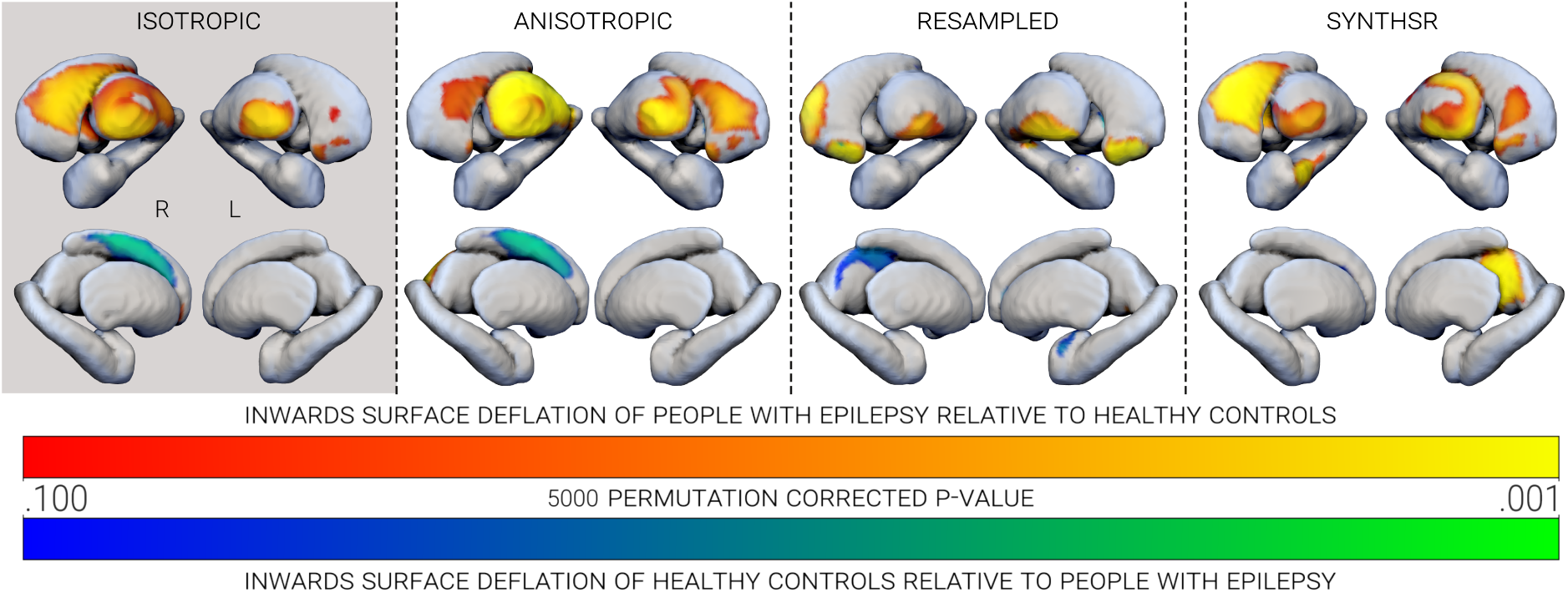
Clusters of subcortical surface deformations between people with IGE relative to healthy controls (warm colours), and vice versa (cool colours), observed across image types. Only clusters with *p corr* < .100 are shown, which were derived from one-sided permutation (*n-perm =* 5000) testing.

**Table 4.**
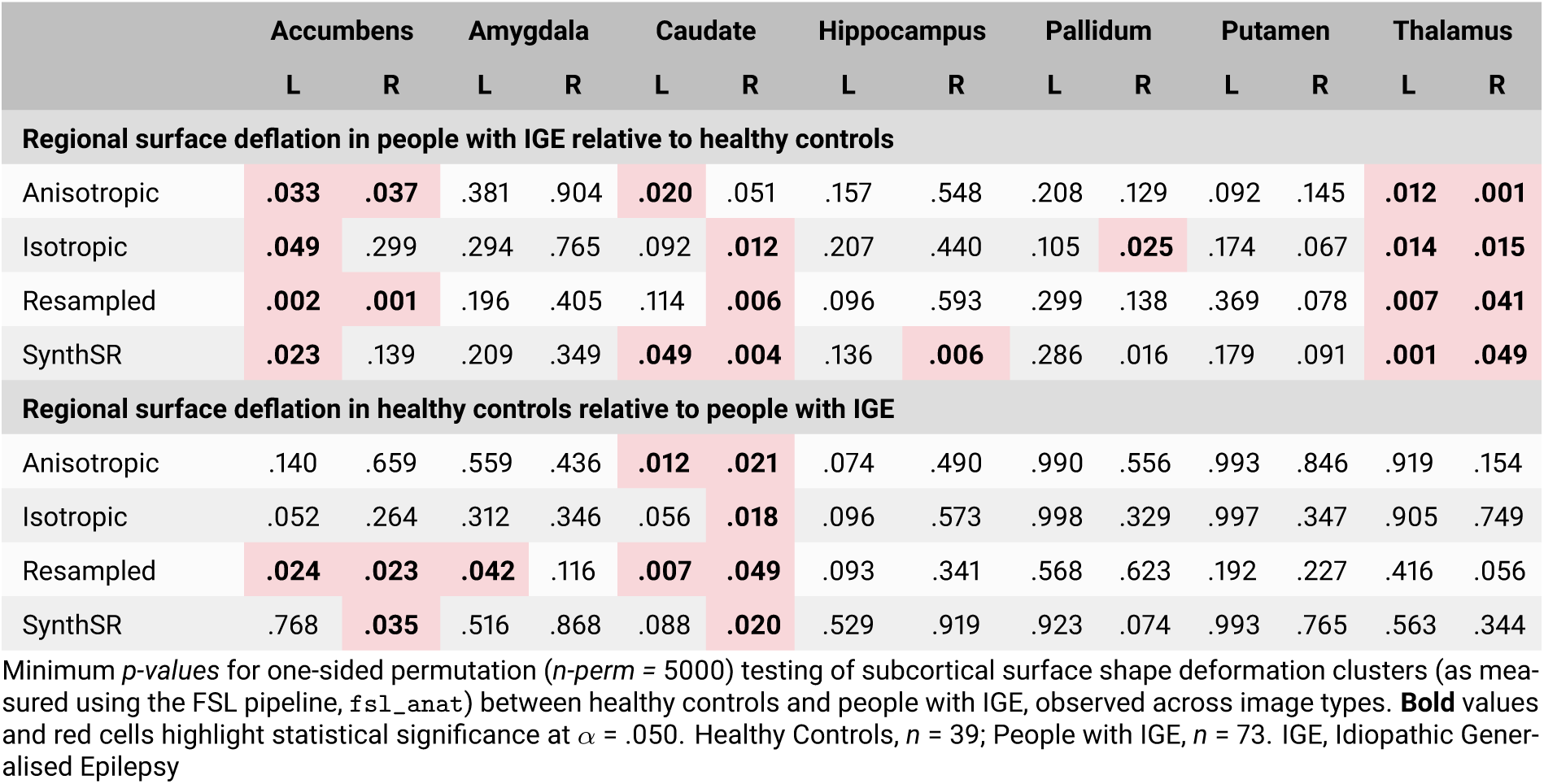
Subcortical surface shape comparisons.

In the pwE, permutation testing of the analogue images consistently identified potential true-positive abnormalities (atrophy of the thalamus bilaterally, atrophy of the left accumbens, and hypertrophy of the right caudate), but failed to detect any deflation of the right pallidum (see table 4 and Figure 5). In addition, based on the 2D-T1 scans regional surface deflation of the right caudate narrowly failed to reach significance (*p corr* = .051), unlike the false-positive deflation in the right accumbens (*p corr* = .037) and the left caudate (*p corr* = .020), or inflation in the left caudate (*p corr* = .012). Despite successfully identifying deflation in the right caudate (*p corr* = .006) of the pwE, comparisons in the res-T1 images also returned several false-positive results: deflation in the right accumbens (*p corr* = .001); inflation of the bilateral accumbens (left *p corr* = .024, right *p corr* = .023); inflation of the left amygdala (*p corr* = .042); and inflation of the right caudate (*p corr* = .007). Similarly, the syn-T1 image comparisons correctly identified inwards surface deflation of the right caudate (*p corr* = .004) in pwE, however erroneously identified deflation of the left caudate (*p corr* = .049) and right hippocampus (*p corr* = .006), and inflation of the right accumbens (*p corr* = .035).

For a supplementary analysis, we also looked at potential subcortical correlates of drug-responsiveness in our cohort. Briefly, based on the 3D-T1 scans, no volumetric abnormalities reached significance; surface shape abnormalities were found, however, between HC and pwIGE. Inwards surface deflation of the pwIGE (relative to HC) was substantiated in the caudate and thalamus bilaterally, as well as the right pallidum and putamen. Outwards surface inflation of the caudate (again, bilaterally) also reached significance. Although not explored further, the full subgroup results are detailed in Appendices 2 and 3.

## 4 Discussion

### 4.1 Imaging summary

This study aimed to evaluate the efficacy of available classical and ML methods for improving on the coherence of quantitative structural metrics estimated from 3D-T1 MRI scans with those from natively anisotropic analogues. We have done so using an anisotropic dataset representative of that which would be typically available retrospectively. Rigorous control during the acquisition of our dataset (2D and 3D scans acquired in the same session and scanner) makes this sample ideal for resolution comparisons, despite its modest size, and the inclusion of HC, pwIGE, and pwDRE provides a means of assessing clinical utility. We supplement this with a preliminary independent evaluation of the new FreeSurfer `recon-all-clinical` pipeline.

Estimates of cortical volume and cortical thickness were markedly reduced in all of the analogue images, although variations in subcortical volume estimates were more evenly dispersed around the ground truth. DSC analyses showed the greatest concordance between the 3D-T1 and 2D-T1 scans, although overlaps were fairly consistent across the cortical parcellations in general—this is likely influenced by the coarseness of our parcellation atlas and decision to coregister the parcellation images. Overlap improved slightly for the subcortical structures, and again, the 2D-T1 scans were the most similar to our ground truth data. In the context of the previous analyses, it is unsurprising that ICC were highest between the metrics extracted from the 3D-T1 and 2D-T1 scans, followed by the res-T1, syn-T1, and lastly the dl-T1 metrics. Summary statistics from the FreeSurfer RA and RC pipelines were generally comparable, although variance in estimates of cortical thickness was markedly higher in the RA pipeline, whereas ICCs and DSCs were slightly reduced.

In the 3D-T1 scan comparisons, pwE presented with evidence of distributed subcortical shape differences indicative of both atrophy and hypertrophy, relative to HC. There is an abundance of evidence for subcortical abnormalities related to the development and maintenance of an epileptogenic environment, which supports the hypothesis that these shape differences are representative of ‘real’ effects. (Billot, Greve, et al., 2023; J. Du et al., 2020; Huang et al., 2017; Zhao et al., 2021) Whilst a subset of shape abnormalities were also identifiable in the 2D-T1 scans (left accumbens, right caudate, and thalamus bilaterally), false positives (right accumbens and left caudate) and false negatives (deflation of the the right caudate and pallidum) add to the scepticism surrounding the reliability of analyses based on data with a low spatial resolution. Greater statistical rigour could indeed be employed to control for false positives, but the possibility of false-negative results might encourage researchers to make use of experimental image synthesis methodologies that are insufficiently validated. In this study, neither resampling nor image synthesis with SynthSR provided evidence for specifically increased sensitivity to true positive effects in data with low spatial resolution, suggesting that even with necessarily increased statistical rigour, evaluation of untransformed data might prove more illuminating.

### 4.2 Clinical summary

With modern analysis techniques, imaging abnormalities in IGE are reported with increasing frequency and evidence suggests that associated structural abnormalities extend beyond putative atrophy, potentially including abnormal orientations or regional subcortical thickening (hypertrophy). (Caciagli et al., 2019; Zubal et al., 2025) Limited by an IGE sample less than a quarter the size of that used for the 2018 ENIGMA study, and the relative heterogeneity of IGE as an epilepsy subtype, we opted to focus on examining subcortical morphometry for the clinical arm of this project. (McKavanagh et al., 2023; Whelan et al., 2018)

A subset of the data analysed in this paper has been examined previously by Brownhill et al. (2021), who found evidence of pallidal, putaminal, and thalamic volumetric reductions (relative to controls) in the truncated IGE cohort. This study recapitulates and expands on the findings of previous structural imaging analyses into the neural correlates of IGE. Firstly by extending the findings of Brownhill et al. (2021) and identifying bilateral pallidal volume reductions, but also by demonstrating that whilst volumetric analyses might lack the necessary sensitivity to identify IGE-specific abnormalities in other key regions, subcortical shape analyses can potentially identify pathology-related focal atrophy and hypertrophy. Specifically, we highlight abnormal inwards surface deflation in the accumbens, caudate, and thalamus (concordant with volumetric abnormalities found in Whelan et al. (2018)), and outwards regional inflation in the caudate-despite the lack of any volumetric abnormalities, such as those specific to the pallidum.

Hypertrophy of the amygdalae, associated with mTLE, is not present, and could potentially influence the distinct cognitive profiles of mTLE and IGE. (Ratcliffe et al., 2020; Zubal et al., 2025) Our findings support the hypothesis that the interaction between structural abnormalities and phenotype in IGE are multifaceted and complex, and that further focused study could deliver insights into the impact of subcortical/projection network reorganisation on seizures presentation and comorbid cognitive impairment. (Rodriguez-Cruces et al., 2022)

IGE is a heterogenous subtype of epilepsy, the phenotypes of which the present sample is insufficient to represent reliably. Nonetheless, our subgroup analyses indicate that structural abnormalities in individuals with drug-responsive IGE were significantly greater than in drug-resistant IGE. So whilst tenuous, our results could imply that regional structural abnormalities of the caudate and thalamus result from, instead of pathology, the mechanism by which seizure control is achieved; ASM-related network disruption could encourage reorganisation of key network hubs in the subcortical structures, preventing ictal spreading. (Royer et al., 2022)

### 4.3 Considerations for the current study

In the context of broader neuroimaging analysis, the scope of this study is limited. Structural imaging is a rapidly advancing technology and the popularity of ML methods is driving accelerated development. It is therefore likely that the methods used in this study will soon be updated, superseded, or made redundant. Whilst this does not impact the validity of our findings, it should be stated that our conclusions cannot be applied *en masse*, and generalisation is at the discretion of the researcher. Compared to some multi-centre studies, our sample is relatively small and in the age of big data the reliability of large-*n* studies is desirable. Nonetheless, a sample of research-quality MRI in 112 participants is reflective of the scope of the average prospective clinical study, and avoids the statistical complications involved in harmonising multi-site data.

Even with an anisotropic acquisition, we have used research quality data with predictable and consistent error. Though we are unable to generalise our findings to other forms of bias (i.e. movement artifacts, contrast-enhancement, reduced field-of-view), we are confident enough in the prevalence of anisotropic clinical MRI data to be assured of the relevance of our findings.

### 4.4 Clinical morphometry

Our subcortical morphometry results indicate that the bias introduced by PVE can manifest as a type-one or type-two error, a false positive or negative respectively (relative to our silver standard 3D-T1 scans, which will themself contain an indefinite but small measure of error). In the volume comparisons, error was reduced in the res-T1 images, whereas the model-based syn-T1 images introduced substantially more false positives. In the subcortical shape analyses, performance was comparable between the analogue inputs, although res-T1 indicated more false positives here—potentially influenced by the relative alignments of the subcortical structures. A tendency to underestimate true relationships is preferable to the alternatives, which suggests that in general, anisotropic clinical imaging data is likely safest to analyse with modern techniques if presented appropriately. Our supplementary clinical analyses (Appendix 2) broadly confirm this, showing that preemptive resampling can introduce large amounts of false positives. The 2D-T1 and syn-T1 images performed comparably, however, making a definite judgement of efficacy unreliable at this time. It is worth noting that despite recommended input guidelines, in this study preprocessing efficacy was relatively unaffected by anisotropy, indicating that 2D-T1 scans are suitable for use with normative methods (like lesion network symptom mapping). (Schaper et al., 2023) Indeed, the res-T1 images were also the most prone to error in the FSL `run_first_all` pipeline. Normative modelling of clinical data is more accessible than ever, with the proliferation of open-source datasets and powerful contemporary imaging tools such as HD-BET, synthstrip, and boundary based registration. (Greve & Fischl, 2009; Hoopes et al., 2022; Isensee et al., 2019) Researchers should consider using clinical MRI as pilot data: the reduced sensitivity of anisotropic data does not fully preclude its utility in establishing trend-level relationships, such as surface deviations of the accumbens and thalamus related to IGE in this study; PVE-based false positives are less common than in resampled data; and there is substantial unexplored imaging data available, especially in rare or difficult to scan clinical cohorts.

### 4.5 Model-based image preprocessing

The model we used with DL+DiReCT was optimised for the purpose of estimating cortical morphometry in contrast-enhanced lesioned imaging data. The performance of DL+DiReCT in this study should not be taken as indicative of its ability to perform this stated task, but is presented instead as another model-based method with which to compare SynthSR, and as a demonstration of the variance that can be introduced through methodological decisions, compounded by inappropriate use of experimental preprocessing steps. Because of this, we have not examined the use of DL+DiReCT when provided with different input images.

Nonetheless, disparity as a (probable) result of inappropriate model application was demonstrated in this study by the overall poor performance of DL+DiReCT. Furthermore, the DiReCT cortical thickness measurements employed in the DL+DiReCT have previously been shown to systematically differ from those provided by FreeSurfer. (Tustison et al., 2014) Whilst DL+DiReCT’s potential application in generalised contrast-enhanced data is promising, it warrants further independent investigation beyond the scope of this paper.

Unlike DL+DiReCT, SynthSR claims applicability to a broad range of use cases under the blanket of generalised contrast and resolution synthesis. (Iglesias et al., 2023) The authors posit that due to its training data, SynthSR is robust to variations in resolution, contrast, and pathology, providing a general algorithm for transforming any clinical MRI into a synthetic 1mm MP-RAGE image. This is supported by its ability to facilitate distinction between the brains of people with Alzheimer’s Disease and healthy controls, or improve the application of preprocessing steps in lesioned images. In this study, we have shown that there are contexts in which SynthSR is not an appropriate solution for image anisotropy—namely in the evaluation of subcortical surface shape. This is possibly a consequence of the specificity of the abnormalities (IGE is typically MR-negative after visual inspection), which causes them to be insufficiently represented in SynthSR’s training data. Furthermore, we have demonstrated that synthetic images do not increase agreement (compared to untransformed 2D images) with the ground truth for the estimation of morphometric properties. Synthetic images represent data that has been fundamentally changed, which is reflected in morphometric analysis, and should be clearly stated throughout dissemination.

It is worth noting the relatively similar performance of SynthSR and DL+DiReCT in several of our comparisons, despite their differing purposes; both models failed to noticeably increase the DSC or ICCs of our metrics relative to the 3D-T1 scans, when compared with the 2D-T1 scans. Indeed, both models typically underperformed when compared to using the untransformed anisotropic data, suggesting that further training is needed—especially in the case of SynthSR, which claims to be generalisable. The salient question to ask is, at this stage, why didn’t the generalisable SynthSR perform better than the (arguably) inappropriate DL+DiReCT?

### 4.6 Resampling

Perhaps unsurprisingly, metrics estimated from the res-T1 images were comparable to those from the 2D-T1 scans. Cortical thickness error was seemingly attenuated through the use of FreeSurfer’s RC pipeline, although mixed results in the other metrics preclude a blanket recommendation of that particular methodology.

One of the more striking findings from this study was the rate of false positives present in the clinical comparisons when using the res-T1 images, which suggests that researchers hoping to upsample anisotropic data (to facilitate examination with software requiring isotropic inputs) may inadvertently bias their data - even beyond exacerbating PVE. We noted registration errors following the use of run_first_all in the res-T1 images (and no others), which undoubtedly contributed to the differences seen in the results.

### 4.7 Machine learning in clinical imaging

The consistent underestimation of grey matter volume and thickness in the analogue images suggests that lower spatial resolution biases PVE towards white matter and extraparenchymal tissue (i.e. the meninges). This is seemingly exacerbated by preprocessing methods, hence the greater disparity between our ground truth and the estimates from the res-T1, syn-T1, and dl-T1 images. This behaviour is difficult to avoid in interpolation, however there is potential for a model-based solution, provided that appropriate data is used when training the model. The choice of training data used when creating am ML model is crucial to the function of the resultant network. For example, in the presence of uncertainty, a super-resolution model trained on healthy data could be expected to attenuate pathology present in input data, making it more difficult to identify despite the improved clarity of the output image. Generally, the data a model is trained on will provide a measure of bias (overfitting reflecting priors) with downstream impacts that make it difficult to quantify, and the variety of the data should be proportional to the breadth of the datasets it is applicable to. (Gambhir et al., 2022; Konell et al., 2024) Caution is therefore advisable when using models claiming input agnosticism, due to the massive variability of neurological pathology.

### 4.8 FreeSurfer Pipelines

By including estimates of cortical and subcortical morphometry from both FreeSurfer’s RA and RC pipelines, we are able to present a systematic, non-exhaustive, evaluation of RC’s efficacy based on multiple types of input images. The differences in cortical thickness variances suggest that the RC pipeline succeeds in reducing the tendency for outlier thickness estimation in anisotropic input data, as is shown in table 1. Nonetheless, there was also some variance between outputs of the two pipelines when provided with the 3D-T1 scans, particularly in the cortical thickness estimates, and ICCs were consistent (or slightly worse) across all metrics. Unaddressed concerns about model-based analyses preclude an unmitigated recommendation of FS (in spite of its increased robustness, and faster processing time), especially as in this study there is no evidence for its increased accuracy (relative to the ground truth) compared to running the standard `recon-all` pipeline on anisotropic data. Furthermore, the two FreeSurfer pipelines, whilst similar, are methodologically distinct, and researchers should be mindful of comparing results between the two. Due to the novelty of the RC pipeline there is likely to be a period of scarcity for external validation, limiting the generalisability of RC outputs. In addition, the ubiquity of results obtained with the FreeSurfer RA pipeline is a strong motivation to opt for it over the RC pipeline, despite the flexibility of the model-based approach relative to the Bayesian methodology. Of course, there is already considerable debate about version control within FreeSurfer, and it has been suggested that morphometry analyses should ideally be repeated with multiple versions of the software, in order to provide a more comprehensive estimate. (Korbmacher et al., 2024) So whilst our results do suggest reasonable consistency between approaches, they also emphasise the trade-off between robustness and accuracy in ML image processing. (Raghunathan et al., 2020) For FreeSurfer’s `recon-all-clinical` pipeline, this trade-off should be explored across multiple datasets, and is beyond the scope of this study to fully evaluate.

### 4.9 Future considerations

As ML methods become increasingly commonplace, it is important to remember general limitations and strengths. Models cannot create data that was not recorded, only provide an estimate, and always with some level of uncertainty. ML methods are not sensitive to inter-subject heterogeneity. And whilst models will only improve over time with techniques like dropout modelling, overfitting will be difficult to avoid entirely. Just as projects/software such as MELD (Spitzer et al., 2022), HD-BET (Isensee et al., 2019), and AMS (Boaro et al., 2022; Kaczmarzyk et al., 2023) have leveraged model-based processing to achieve greater tissue classification accuracy than was previously possible, researchers should be aware of the reduction in interpretability between traditional classifiers (like those mentioned) and deep learning algorithms designed to extract hidden features. (Yuan et al., 2022) Whilst it is tempting to adopt the general consensus that machine learning will revolutionise information processing, at this early stage a measure of scepticism should be employed when working with ML methods, including implementing manual checks whenever possible and encouraging methodological scrutiny.

### 4.10 Conclusion

Any of the methods presented herein may be readily employed in an attempt to coerce morphometric information from low-resolution (retrospective or clinical) imaging data. However, we have shown that contrary to popular opinion, the evaluation of 2D-T1 data may constitute the most appropriate strategy, when possible. In a dataset typical of a clinical context, analogue images provided by resampling and synthesis techniques showed a reduction in the relative preservation of morphometric properties otherwise discernible from 2D-T1 scans. Neuroimaging researchers should carefully evaluate their choice of preprocessing before proposing morphometric associations, and when applicable, clearly state that results are based on manipulated data.

## Data Availability

Data available on request from the authors. Code freely available on GitHub.

https://github.com/C-Ratcliffe/221216_Proj-IS

## Abbreviations

ANTs: Advanced Normalization Tools
ASM: Anti-seizure Medication
CNN: Convolutional Neural Network
DSC: Dice Similarity Coefficients
dl-T1: DL+DiReCT Outputs
FSL: FMRIB Software Library
GLM(s): General Linear Model(s)
HC: Healthy Controls
ICC: Intraclass Correlation Coefficients
IGE: Idiopathic Generalised Epilepsies
ML: Machine Learning
PVE: Partial Volume Effects
pwDRE: People with Drug-resistant idiopathic generalised Epilepsy
pwE: People with idiopathic generalised Epilepsy
pwIGE: People with drug-responsive Idiopathic Generalised Epilepsy
RA: `recon-all`
RC: `recon-all-clinical`
res-T1: Resampled isotropic T1
syn-T1: SynthSR isotropic T1 images
TPV: Total Parenchymal Volume
2D-T1: Anisotropic
3D-T1: Isotropic

## Data and code availability

Data available on request from the authors. Code freely available on GitHub.

## Author contributions

CR: Conceptualisation, investigation, visualisation, writing - original draft preparation, writing - review and editing.

PNT: Writing - review and editing.

CdB: Conceptualisation.

KD: Resources.

SB: Resources.

AM: Writing - review and editing.

SSK: Writing - review and editing, supervision.

## Funding

SK acknowledges support from the UK Medical Research Council (Grant Number MR/S00355X/1).

## Declaration of competing interests

The authors declare no conflicts of interest. We confirm that we have read the Journal’s position on issues involved in ethical publication and affirm that this report is consistent with those guidelines.

## Supplementary material

Scripts: GitHub

## Appendix

## Appendix 1

**Table a1.** Sample demographics.

| Study ID | Sex | Age at Scan | Group | Total Parenchymal Volume (mm<sup>3</sup>) | Study ID | Sex | Age at Scan | Group | Total Parenchymal Volume (mm<sup>3</sup>) |
| --- | --- | --- | --- | --- | --- | --- | --- | --- | --- |
| sub-001 | F | 31 – 35 | pwIGE | 1500325.74 | sub-057 | F | 41 – 45 | HC | 1549914.99 |
| sub-002 | F | 21 – 25 | pwIGE | 1542291.40 | sub-058 | F | 16 – 20 | pwDRE | 1695651.18 |
| sub-003 | M | 16 – 20 | pwIGE | 1861860.90 | sub-059 | M | 16 – 20 | pwDRE | 1734680.68 |
| sub-004 | F | 16 – 20 | pwIGE | 1396310.05 | sub-060 | F | 31 – 35 | pwDRE | 1301169.69 |
| sub-005 | M | 21 – 25 | pwIGE | 1637994.16 | sub-061 | F | 21 – 25 | HC | 1472779.06 |
| sub-006 | F | 56 – 60 | pwIGE | 1493748.84 | sub-062 | M | 51 – 55 | pwDRE | 1499155.29 |
| sub-007 | M | 21 – 25 | pwIGE | 1549106.18 | sub-063 | F | 31 – 35 | HC | 1497035.28 |
| sub-008 | F | 21 – 25 | pwIGE | 1587915.52 | sub-064 | F | 31 – 35 | HC | 1431230.78 |
| sub-009 | F | 31 – 35 | pwIGE | 1588225.40 | sub-065 | F | 31 – 35 | HC | 1454569.74 |
| sub-010 | M | 36 – 40 | pwIGE | 1511318.51 | sub-066 | F | 26 – 30 | pwDRE | 1680187.58 |
| sub-011 | M | 66 – 70 | pwIGE | 1533209.66 | sub-067 | M | 36 – 40 | HC | 1927143.97 |
| sub-012 | F | 46 – 50 | pwIGE | 1495030.25 | sub-068 | M | 31 – 35 | HC | 1562383.85 |
| sub-013 | M | 16 – 20 | pwIGE | 1953398.17 | sub-069 | F | 41 – 45 | HC | 1466323.57 |
| sub-014 | F | 21 – 25 | pwIGE | 1460892.16 | sub-070 | F | 21 – 25 | pwDRE | 1369512.9 |
| sub-015 | M | 31 – 35 | pwIGE | 1653925.05 | sub-071 | M | 26 – 30 | HC | 1713088.98 |
| sub-016 | M | 16 – 20 | pwIGE | 1564495.70 | sub-072 | F | 31 – 35 | pwDRE | 1490116.37 |
| sub-017 | M | 36 – 40 | pwIGE | 1423657.57 | sub-073 | M | 36 – 40 | HC | 1722978.46 |
| sub-018 | F | 21 – 25 | pwIGE | 1995978.41 | sub-074 | F | 26 – 30 | HC | 1531661.85 |
| sub-019 | M | 21 – 25 | pwIGE | 1726997.75 | sub-075 | F | 31 – 35 | HC | 1452227.41 |
| sub-020 | F | 36 – 40 | pwIGE | 1755597.02 | sub-076 | F | 26 – 30 | pwDRE | 1428725.09 |
| sub-021 | F | 31 – 35 | pwIGE | 1522907.27 | sub-077 | M | 41 – 45 | HC | 1658457.47 |
| sub-022 | F | 31 – 35 | pwIGE | 1529185.05 | sub-078 | F | 41 – 45 | pwDRE | 1259660.17 |
| sub-023 | M | 21 – 25 | pwIGE | 1586201.15 | sub-079 | M | 16 – 20 | pwDRE | 1804608.9 |
| sub-024 | F | 16 – 20 | pwIGE | 1740328.94 | sub-080 | M | 21 – 25 | HC | 1824748.21 |
| sub-025 | F | 56 – 60 | pwIGE | 1473819.61 | sub-081 | F | 41 – 45 | pwDRE | 1408431.19 |
| sub-026 | F | 16 – 20 | pwIGE | 1405915.12 | sub-082 | M | 36 – 40 | HC | 1740562.68 |
| sub-027 | M | 21 – 25 | pwIGE | 1816224.09 | sub-083 | F | 21 – 25 | pwDRE | 1505731.72 |
| sub-028 | M | 21 – 25 | pwIGE | 1656447.61 | sub-084 | M | 36 – 40 | pwDRE | 1783313.46 |
| sub-029 | F | 56 – 60 | pwIGE | 1690206.23 | sub-085 | F | 26 – 30 | pwDRE | 1356596.93 |
| sub-030 | F | 56 – 60 | pwIGE | 1391252.34 | sub-086 | F | 36 – 40 | pwDRE | 1562572.83 |
| sub-031 | M | 31 – 35 | pwIGE | 1395181.14 | sub-087 | M | 36 – 40 | pwDRE | 1496065.78 |
| sub-032 | M | 21 – 25 | HC | 1756746.92 | sub-088 | F | 36 – 40 | HC | 1550714.93 |
| sub-033 | M | 21 – 25 | HC | 1632795.68 | sub-089 | M | 21 – 25 | pwDRE | 1538654.83 |
| sub-034 | M | 21 – 25 | HC | 1628394.95 | sub-090 | F | 21 – 25 | pwDRE | 1733437.92 |
| sub-035 | F | 61 – 65 | pwDRE | 1559574.71 | sub-091 | M | 16 – 20 | pwDRE | 1916703.41 |
| sub-036 | M | 21 – 25 | HC | 1700479.91 | sub-092 | F | 26 – 30 | HC | 1570408.41 |
| sub-037 | M | 26 – 30 | pwDRE | 1686863.59 | sub-093 | F | 26 – 30 | pwDRE | 1412396.84 |
| sub-038 | F | 26 – 30 | HC | 1724797.87 | sub-094 | M | 26 – 30 | pwDRE | 1932541.28 |
| sub-039 | M | 36 – 40 | HC | 1696736.56 | sub-095 | F | 26 – 30 | HC | 1491109.73 |
| sub-040 | M | 21 – 25 | pwDRE | 1506507.38 | sub-096 | F | 31 – 35 | HC | 1397883.2 |
| sub-041 | F | 21 – 25 | HC | 1443795.57 | sub-097 | M | 36 – 40 | HC | 1718302.02 |
| sub-042 | F | 56 – 60 | HC | 1433847.43 | sub-098 | F | 26 – 30 | HC | 1599759.26 |
| sub-043 | M | 21 – 25 | pwDRE | 1799045.87 | sub-099 | M | 31 – 35 | HC | 1768257.64 |
| sub-044 | F | 16 – 20 | pwDRE | 1404680.91 | sub-100 | F | 21 – 25 | HC | 1545723.81 |
| sub-045 | F | 26 – 30 | pwDRE | 1646535.98 | sub-101 | F | 21 – 25 | HC | 1456575.91 |
| sub-046 | F | 26 – 30 | HC | 1587856.12 | sub-102 | M | 26 – 30 | pwDRE | 1559425.3 |
| sub-047 | F | 41 – 45 | pwDRE | 1555237.06 | sub-103 | F | 46 – 50 | pwDRE | 1402154.62 |
| sub-048 | F | 46 – 50 | HC | 1454538.28 | sub-104 | F | 36 – 40 | pwDRE | 1396666.03 |
| sub-049 | M | 41 – 45 | pwDRE | 1741355.94 | sub-105 | M | 31 – 35 | pwDRE | 1513924.64 |
| sub-050 | F | 51 – 55 | pwDRE | 1326614.89 | sub-106 | F | 21 – 25 | HC | 1629950.58 |
| sub-051 | M | 31 – 35 | HC | 1413972.33 | sub-107 | F | 16 – 20 | pwDRE | 1429645.26 |
| sub-052 | F | 36 – 40 | pwDRE | 1582578.7 | sub-108 | F | 21 – 25 | pwDRE | 1341604.36 |
| sub-053 | M | 46 – 50 | pwDRE | 1876719.97 | sub-109 | F | 21 – 25 | pwDRE | 1520121.37 |
| sub-054 | F | 36 – 40 | HC | 1722298.96 | sub-110 | M | 21 – 25 | pwDRE | 1683436.33 |
| sub-055 | F | 36 – 40 | pwDRE | 1326222.36 | sub-111 | M | 41 – 45 | HC | 1480935.45 |
| sub-056 | F | 26 – 30 | HC | 1352827.48 | sub-112 | M | 26 – 30 | pwDRE | 1673906.88 |

Total sample demographics. F, Female; HC, Healthy Controls; M, Male; pwDRE, people with Drug-resistant idiopathic generalised Epilepsy; pwDRE, people with drug-responsive Idiopathic Generalised Epilepsy

## Appendix 2

**Table a2.** Wholebrain descriptive summary statistics.

|  | Wholebrain |  |  |
| --- | --- | --- | --- |
|  | Similarity | Volume | Volume |
|  | (DSC) | (z-score) | (ICC) |
| **Anisotropic** |  |  |  |
| recon-all | 0.94 | -0.13 | 0.97 |
| recon-all-clinical | 0.91 | -0.81 | 0.98 |
| **DL+DiReCT** |  |  |  |
| DeepSCAN | 0.84 | -2.72 | 0.93 |
| **Isotropic** |  |  |  |
| recon-all | Reference |  |  |
| recon-all-clinical | 0.93 | -0.89 | 0.99 |
| **Resampled** |  |  |  |
| recon-all | 0.94 | -0.13 | 0.97 |
| recon-all-clinical | 0.91 | -0.79 | 0.99 |
| **SynthSR** |  |  |  |
| recon-all | 0.94 | 0.32 | 0.98 |

Structural measurements for different image types, extracted from FreeSurfer `recon-all`, FreeSurfer `recon-all-clinical`, and DeepSCAN parcellations. The isotropic scans, anisotropic scans, and resampled images were processed with both FreeSurfer pipelines, the SynthSR-derived images were processed with `recon-all` only, and DeepSCAN was applied to the resampled images as part of the DL+DiReCT pipeline. Z-scores were computed relative to the distribution from the isotropic data, and ICCs, DSCs, and z-scores were computed based on wholebrain segmentation masks. DSC, Dice Similarity Coefficient; ICC, Intraclass Correlation Coefficient

**Table a3.**
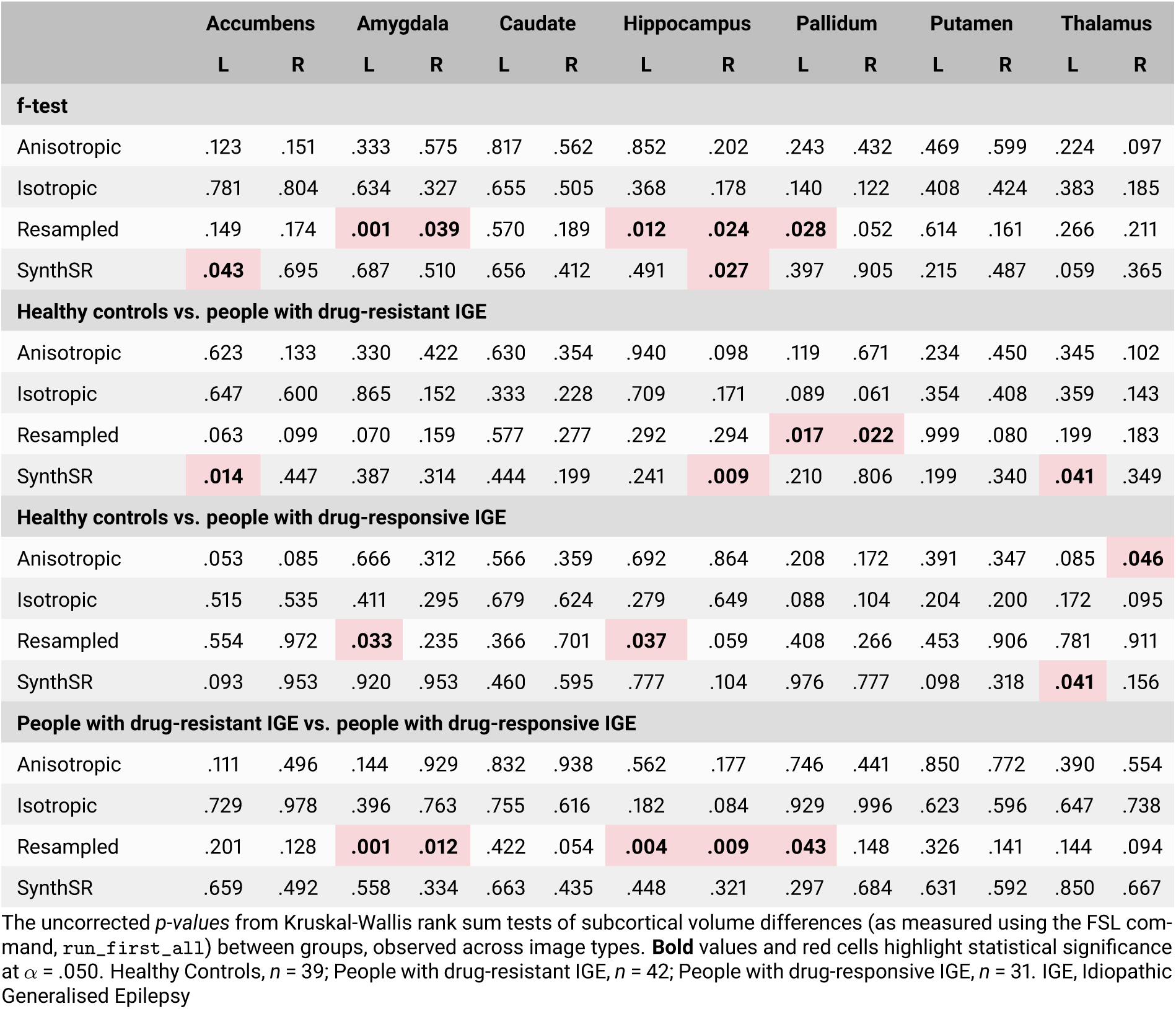
Subcortical volume comparisons.

**Table a4.**
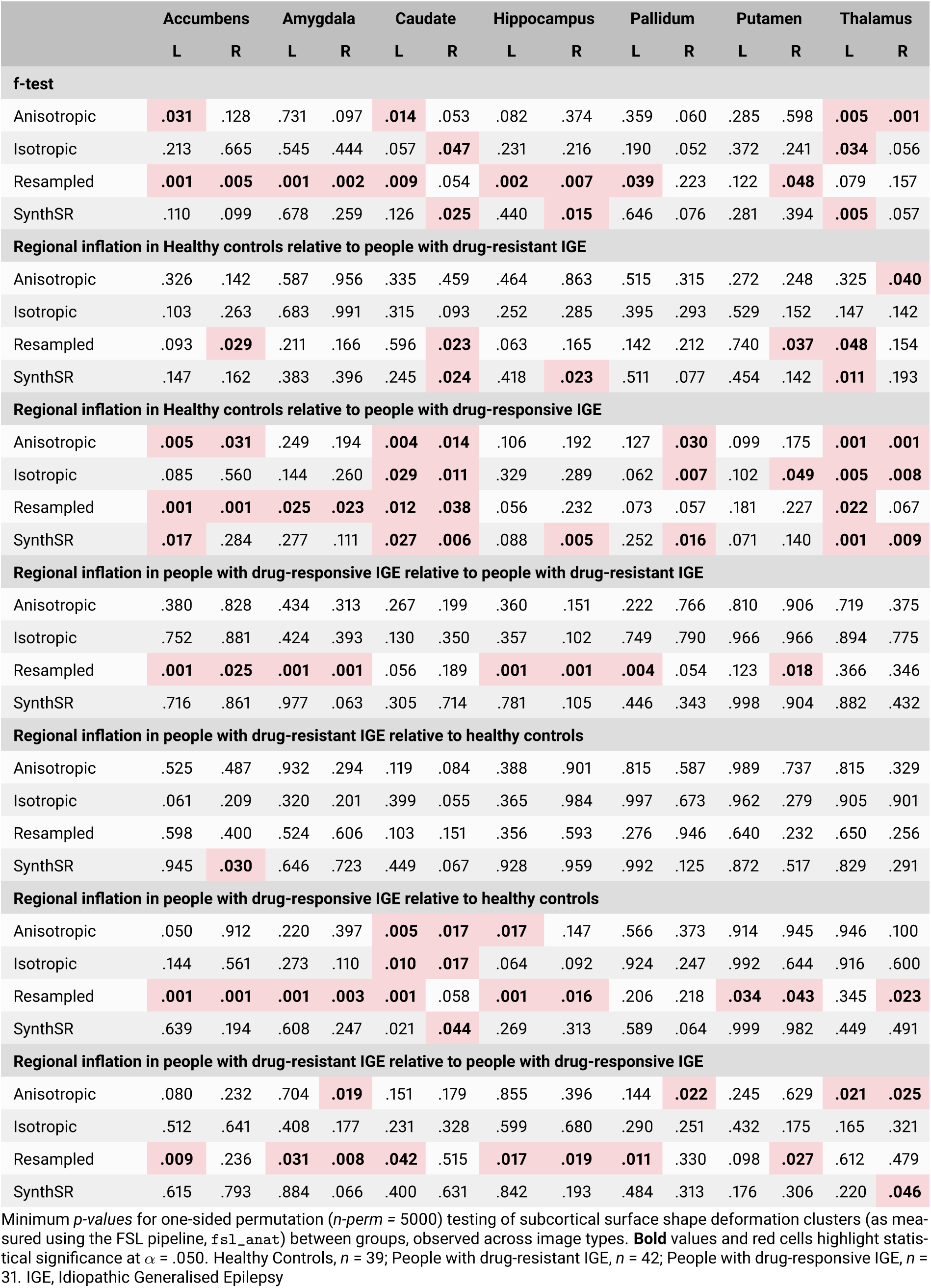
Subcortical surface shape comparisons.

## Appendix 3

**Figure a1.**
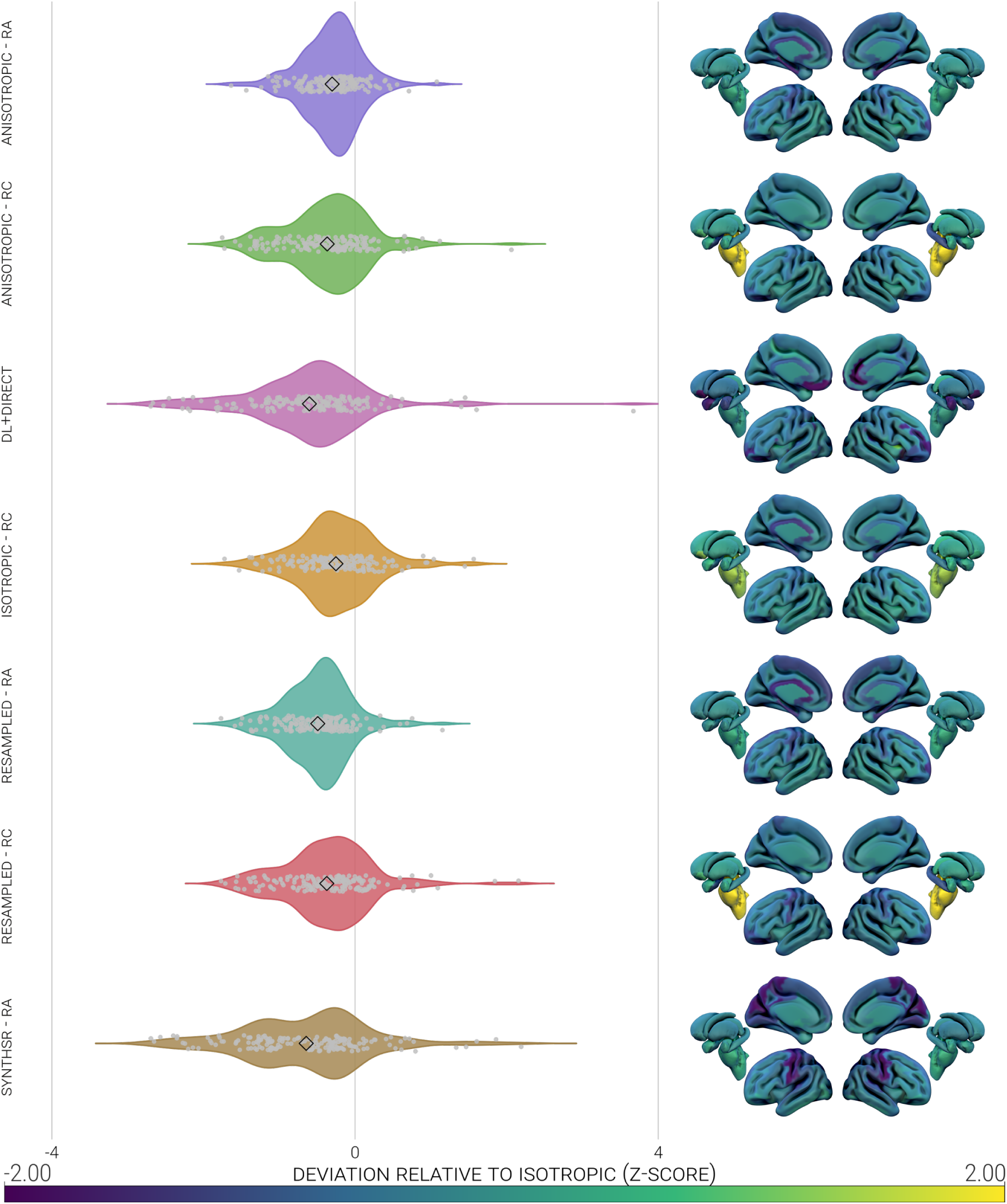
Heatmap and violin plot representations of the volume estimation variance (presented on the scale of -4 to +4 SD) of 174 parcellated regions created from isotropic scans and the various analogues. Regional volume estimates were first standardised to the distribution of the same region in the isotropic data. Regional data were averaged across all participants in each image type, to leave one datum per region per analogue. RA, recon-all; RC, recon-all-clinical

**Figure a2.**
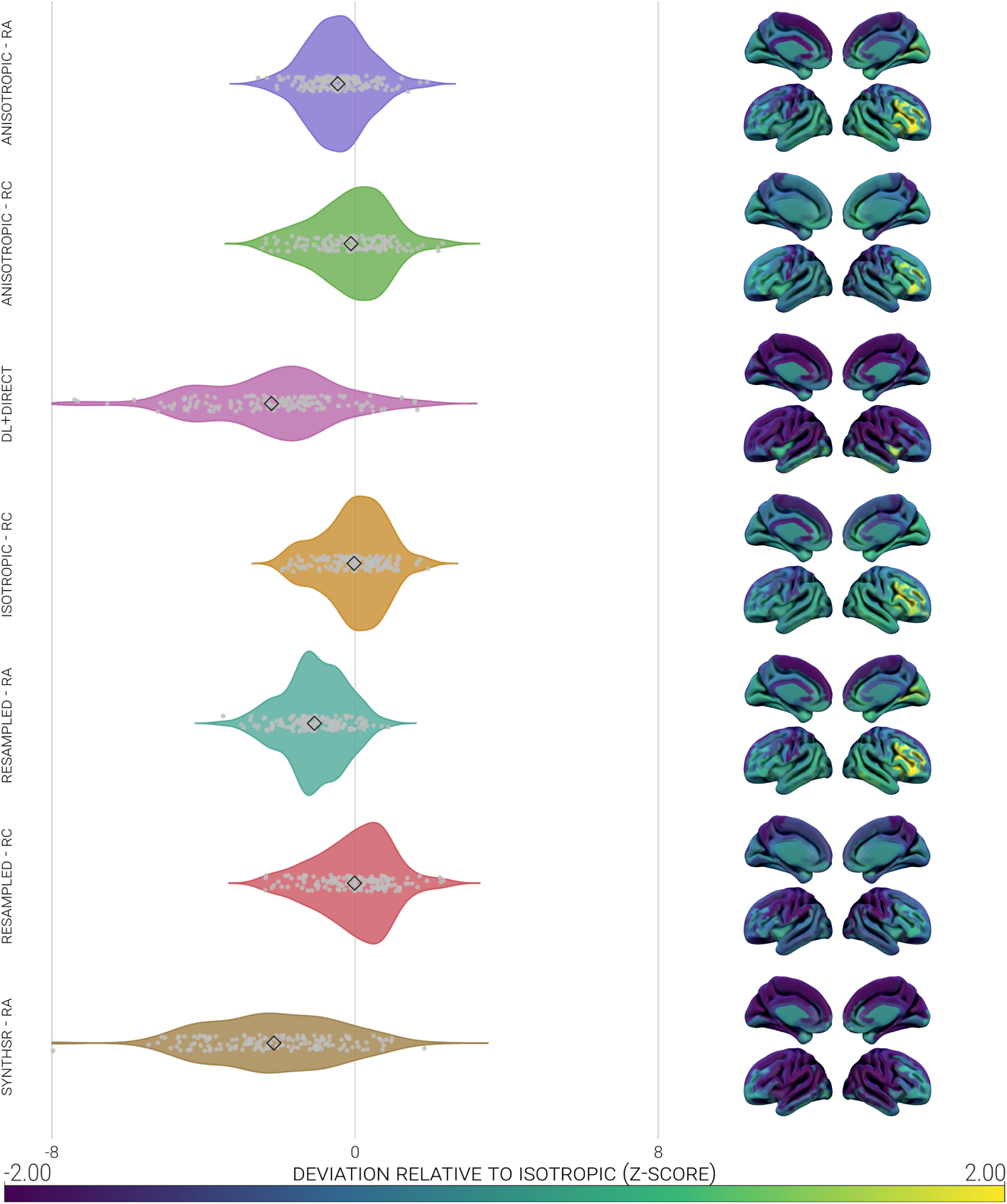
Heatmap and violin plot representations of the thickness estimation variance (presented on the scale of -8 to +8 SD) of 148 parcellated regions created from isotropic scans and the various analogues. Regional thickness estimates were first standardised to the distribution of the same region in the isotropic data. Regional data were averaged across all participants in each image type, to leave one datum per region per analogue. RA, recon-all; RC, recon-all-clinical

**Figure a3.**
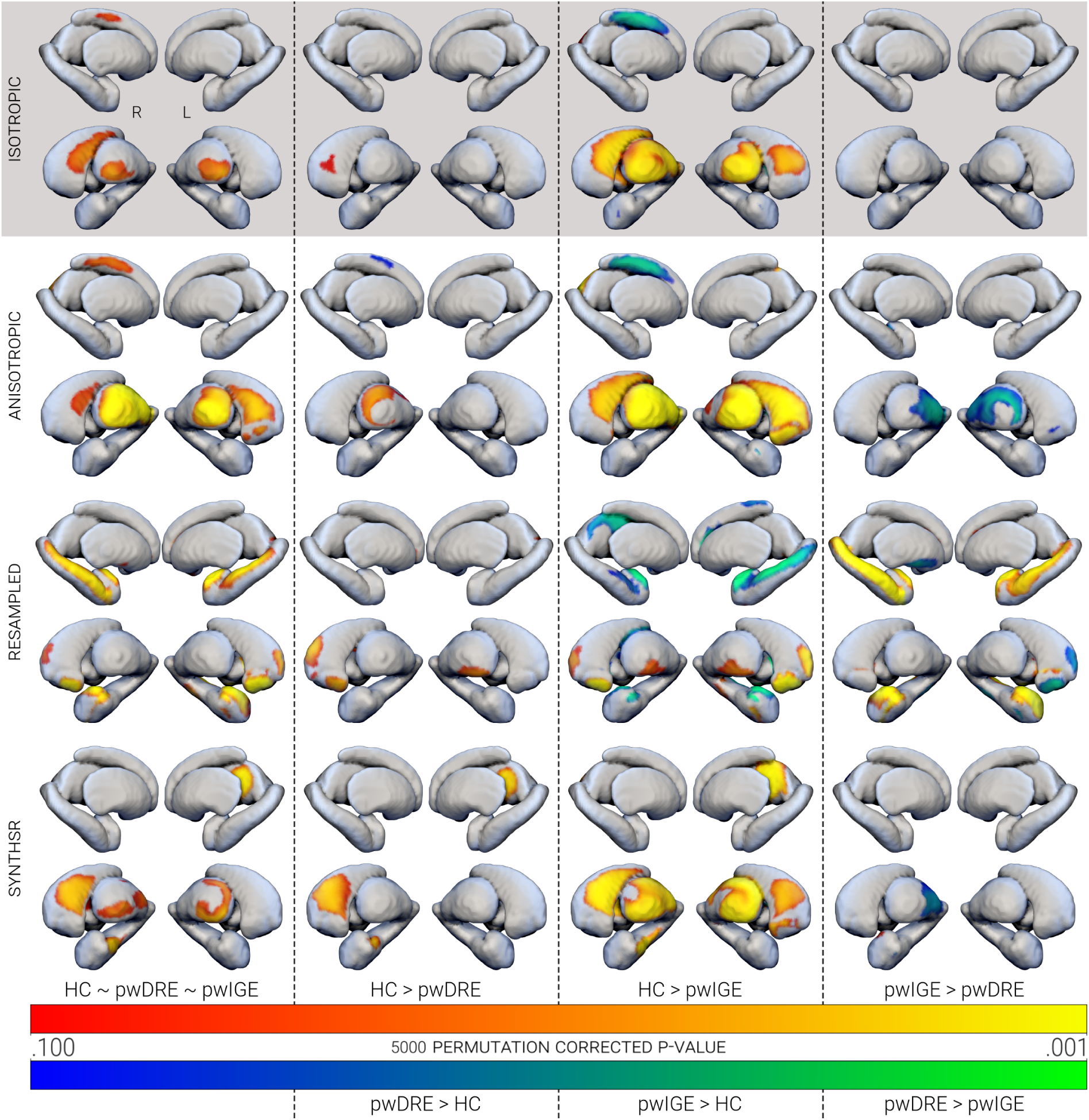
Clusters of subcortical surface shape deformations between people with drug-resistant IGE, people with drug-responsive IGE, and healthy controls, observed across image types. Only clusters with *p corr* < .100 are shown, which were derived with one-sided permutation (*n-perm =* 5000) testing. The direction of the contrast is presented next to the relevant colourbar. HC, Healthy Controls; IGE, Idiopathic Generalised Epilepsy; pwDRE, people with drug-resistant IGE; pwIGE, people with drug-responsive IGE

## Appendix 4

**0.1 Appendix 4.a.**

R environment

- R (R Core Team, 2023)
- RStudio (RStudio Team, 2020)

**0.2 Appendix 4.b.**

R packages

- car (Fox et al., 2024)
- forcats (Wickham & RStudio, 2023)
- ggplot2 (Wickham, 2016)
- ggpubr (Kassambara, 2025)
- MiscTools (Henningsen & Toomet, 2019)
- psych (Revelle, 2023)
- RNifti (Clayden et al., 2023)
- svglite (Wickham, Henry, et al., 2023)
- tidyr (Wickham, Vaughan, & Girlich, 2023)

**0.3 Appendix 4.c.**

Visualisation software

- GIMP (The GIMP Development Team, 2019)
- Inkscape (Inkscape Project, 2020)
- MRICroGL (Rorden & Brett, 2000)
- Scribus (The Scribus Team, 2023)
- Surf Ice (Rorden, 2021)

**0.4 Appendix 4.d.**

Image analysis software

- ANTs (Avants et al., 2009)
- FMRIB Software Library (Jenkinson et al., 2012)
- FreeSurfer (Fischl, 2012)
- Nibabel (Brett et al., 2023)
- DL+DiReCT (McKinley et al., 2021; Rebsamen et al., 2020, 2022)
- SynthSR (Billot, Greve, et al., 2023; Iglesias et al., 2023)

## Notes

### Competing Interest Statement

The authors have declared no competing interest.

### Funding Statement

SSK acknowledges support from the UK Medical Research Council (Grant Number MR/S00355X/1).

### Author Declarations

North West - Liverpool Central Research Ethics Committee gave ethical approval/favourable opinion for this work. REC reference 14/NW/0332).

### Summary of Updates

Following reviewer comments, the paper underwent substantial revisions, including the addition of another 42 cases (people with drug-resistant idiopathic generalised epilepsy) and a new processing arm (FreeSurfer's recon-all-clinical pipeline). 70% of the text, and all of the figures, have been overhauled.

## References

Avants, B. B., Tustison, N., & Johnson, H. (2009). Advanced Normalization Tools (ANTS). Insight J, 2(365), 1–35.

Beghi, E. (2020). The Epidemiology of Epilepsy. Neuroepidemiology, 54(2), 185–191. 10.1159/000503831

Billot, B., Greve, D. N., Puonti, O., Thielscher, A., Van Leemput, K., Fischl, B., Dalca, A. V., & Iglesias, J. E. (2023). SynthSeg: Segmentation of brain MRI scans of any contrast and resolution without retraining. Medical Image Analysis, 86, 102789. 10.1016/j.media.2023.102789

Billot, B., Magdamo, C., Cheng, Y., Arnold, S. E., Das, S., & Iglesias, J. E. (2023). Robust machine learning segmentation for large-scale analysis of heterogeneous clinical brain MRI datasets. Proceedings of the National Academy of Sciences of the United States of America, 120(9), e2216399120. 10.1073/pnas.2216399120

Boaro, A., Kaczmarzyk, J. R., Kavouridis, V. K., Harary, M., Mammi, M., Dawood, H., Shea, A., Cho, E. Y., Juvekar, P., Noh, T., Rana, A., Ghosh, S., & Arnaout, O. (2022). Deep neural networks allow expert-level brain meningioma segmentation and present potential for improvement of clinical practice. Scientific Reports, 12, 15462. 10.1038/s41598-022-19356-5

Bonilha, L., & Keller, S. S. (2015). Quantitative MRI in refractory temporal lobe epilepsy: Relationship with surgical outcomes. Quantitative Imaging in Medicine and Surgery, 5(2), 204–224. 10.3978/j.issn.2223-4292.2015.01.01

Brett, M., Markiewicz, C. J., Hanke, M., Côté, M.-A., Cipollini, B., McCarthy, P., Jarecka, D., Cheng, C. P., Halchenko, Y. O., Cottaar, M., Larson, E., Ghosh, S., Wassermann, D., Gerhard, S., Lee, G. R., Baratz, Z., Wang, H.-T., Kastman, E., Kaczmarzyk, J., . . . freec84. (2023, April). Nipy/nibabel: 5.1.0. 10.5281/zenodo.7795644

Brownhill, D., Chen, Y., Kreilkamp, B. A. K., de Bezenac, C., Denby, C., Bracewell, M., Biswas, S., Das, K., Marson, A. G., & Keller, S. S. (2021). Automated subcortical volume estimation from 2D MRI in epilepsy and implications for clinical trials. Neuroradiology. 10.1007/s00234-021-02811-x

Caciagli, L., Wandschneider, B., Xiao, F., Vollmar, C., Centeno, M., Vos, S. B., Trimmel, K., Sidhu, M. K., Thompson, P. J., Winston, G. P., Duncan, J. S., & Koepp, M. J. (2019). Abnormal hippocampal structure and function in juvenile myoclonic epilepsy and unaffected siblings. Brain, 142(9), 2670– 2687. 10.1093/brain/awz215

Choi, R. Y., Coyner, A. S., Kalpathy-Cramer, J., Chiang, M. F., & Campbell, J. P. (2020). Introduction to Machine Learning, Neural Networks, and Deep Learning. Translational Vision Science & Technology, 9(2), 14. 10.1167/tvst.9.2.14

Clayden, J., Cox, B., Jenkinson, M., Hall, M., Reynolds, R., Fissell, K., Gailly, J.-l., & Adler, M. (2023). RNifti: Fast R and C++ Access to NIfTI Images.

Currie, G., Hawk, K. E., Rohren, E., Vial, A., & Klein, R. (2019). Machine Learning and Deep Learning in Medical Imaging: Intelligent Imaging. Journal of Medical Imaging and Radiation Sciences, 50(4), 477–487. 10.1016/j.jmir.2019.09.005

Das, S. R., Avants, B. B., Grossman, M., & Gee, J. C. (2009). Registration based cortical thickness measurement. NeuroImage, 45(3), 867–879. 10.1016/j.neuroimage.2008.12.016

Delannoy, Q., Pham, C.-H., Cazorla, C., Tor-Díez, C., Dollé, G., Meunier, H., Bednarek, N., Fablet, R., Passat, N., & Rousseau, F. (2020). SegSRGAN: Super-resolution and segmentation using generative adversarial networks — Application to neonatal brain MRI. Computers in Biology and Medicine, 120, 103755. 10.1016/j.compbiomed.2020.103755

Despotović, I., Goossens, B., & Philips, W. (2015). MRI Segmentation of the Human Brain: Challenges, Methods, and Applications. Computational and Mathematical Methods in Medicine, 2015, 450341. 10.1155/2015/450341

Destrieux, C., Fischl, B., Dale, A., & Halgren, E. (2010). Automatic parcellation of human cortical gyri and sulci using standard anatomical nomenclature. NeuroImage, 53(1), 1–15. 10.1016/j.neuroimage.2010.06.010

Du, H., Zhang, Y., Xie, B., Wu, N., Wu, G., Wang, J., Jiang, T., & Feng, H. (2011). Regional atrophy of the basal ganglia and thalamus in idiopathic generalized epilepsy. Journal of Magnetic Resonance Imaging, 33(4), 817–821. 10.1002/jmri.22416

Du, J., He, Z., Wang, L., Gholipour, A., Zhou, Z., Chen, D., & Jia, Y. (2020). Super-resolution reconstruction of single anisotropic 3D MR images using residual convolutional neural network. Neurocomputing, 392, 209–220. 10.1016/j.neucom.2018.10.102

Edelman, R. R., Dunkle, E., Koktzoglou, I., Griffin, A., Russell, E. J., Ankenbrandt, W., Ragin, A., & Carrillo, A. (2009). Rapid whole-brain magnetic resonance imaging with isotropic resolution at 3 Tesla. Investigative Radiology, 44(1), 54–59. 10.1097/RLI.0b013e31818eee3c

Firbank, M. J., Coulthard, A., Harrison, R. M., & Williams, E. D. (1999). Partial volume effects in MRI studies of multiple sclerosis. Magnetic Resonance Imaging, 17(4), 593–601. 10.1016/ S0730-725X(98)00210-0

Fischl, B. (2012). FreeSurfer. NeuroImage, 62(2), 774–781. 10.1016/j.neuroimage.2012.01.021

Fiscone, C., Curti, N., Ceccarelli, M., Remondini, D., Testa, C., Lodi, R., Tonon, C., Manners, D. N., & Castellani, G. (2024). Generalizing the Enhanced-Deep-Super-Resolution neural network to brain MR images: A retrospective study on the Cam-CAN dataset. eNeuro, ENEURO.0458–22.2023. 10.1523/ENEURO.0458-22.2023

Fox, J., Weisberg, S., Price, B., Adler, D., Bates, D., Baud-Bovy, G., Bolker, B., Ellison, S., Firth, D., Friendly, M., Gorjanc, G., Graves, S., Heiberger, R., Krivitsky, P., Laboissiere, R., Maechler, M., Monette, G., Murdoch, D., Nilsson, H., . . . R-Core. (2024, September). Car: Companion to Applied Regression. Gambhir, R., Nachman, B., & Thaler, J. (2022). Bias and priors in machine learning calibrations for high energy physics. Physical Review D, 106(3), 036011. 10.1103/PhysRevD.106.036011

Gopinath, K., Greve, D. N., Das, S., Arnold, S., Magdamo, C., & Iglesias, J. E. (2023, May). Cortical analysis of heterogeneous clinical brain MRI scans for large-scale neuroimaging studies. 10.48550/arXiv.2305.01827

Greve, D. N., & Fischl, B. (2009). Accurate and robust brain image alignment using boundary-based registration. NeuroImage, 48(1), 63–72. 10.1016/j.neuroimage.2009.06.060

Haller, S., Falkovskiy, P., Meuli, R., Thiran, J.-P., Krueger, G., Lovblad, K.-O., Kober, T., Roche, A., & Marechal, B. (2016). Basic MR sequence parameters systematically bias automated brain volume estimation. Neuroradiology, 58(11), 1153–1160. 10.1007/s00234-016-1737-3

Hatton, S. N., Huynh, K. H., Bonilha, L., Abela, E., Alhusaini, S., Altmann, A., Alvim, M. K. M., Balachandra, A. R., Bartolini, E., Bender, B., Bernasconi, N., Bernasconi, A., Bernhardt, B., Bargallo, N., Caldairou, B., Caligiuri, M. E., Carr, S. J. A., Cavalleri, G. L., Cendes, F., . . . McDonald, C. R. (2020). White matter abnormalities across different epilepsy syndromes in adults: An ENIGMA-Epilepsy study. Brain: A Journal of Neurology, 143(8), 2454–2473. 10.1093/brain/awaa200

Henningsen, A., & Toomet, O. (2019). Misctools: Miscellaneous Tools and Utilities.

Hoopes, A., Mora, J. S., Dalca, A. V., Fischl, B., & Hoffmann, M. (2022). SynthStrip: Skull-stripping for any brain image. NeuroImage, 260, 119474. 10.1016/j.neuroimage.2022.119474

Huang, Y., Shao, L., & Frangi, A. F. (2017). Simultaneous Super-Resolution and Cross-Modality Synthesis of 3D Medical Images Using Weakly-Supervised Joint Convolutional Sparse Coding. Proceedings of the IEEE Conference on Computer Vision and Pattern Recognition, 6070–6079.

Iglesias, J. E., Billot, B., Balbastre, Y., Magdamo, C., Arnold, S. E., Das, S., Edlow, B. L., Alexander, D. C., Golland, P., & Fischl, B. (2023). SynthSR: A public AI tool to turn heterogeneous clinical brain scans into high-resolution T1-weighted images for 3D morphometry. Science Advances, 9(5), eadd3607. 10.1126/sciadv.add3607

Iglesias, J. E., Billot, B., Balbastre, Y., Tabari, A., Conklin, J., Gilberto González, R., Alexander, D. C., Golland, P., Edlow, B. L., & Fischl, B. (2021). Joint super-resolution and synthesis of 1 mm isotropic MP-RAGE volumes from clinical MRI exams with scans of different orientation, resolution and contrast. NeuroImage, 237, 118206. 10.1016/j.neuroimage.2021.118206

Inkscape Project. (2020, April). Inkscape.

Isensee, F., Schell, M., Pflueger, I., Brugnara, G., Bonekamp, D., Neuberger, U., Wick, A., Schlemmer, H.-P., Heiland, S., Wick, W., Bendszus, M., Maier-Hein, K. H., & Kickingereder, P. (2019). Automated brain extraction of multisequence MRI using artificial neural networks. Human Brain Mapping, 40(17), 4952–4964. 10.1002/hbm.24750

Jenkinson, M., & Smith, S. (2001). A global optimisation method for robust affine registration of brain images. Medical Image Analysis, 5(2), 143–156. 10.1016/s1361-8415(01)00036-6

Jenkinson, M., Bannister, P., Brady, M., & Smith, S. (2002). Improved optimization for the robust and accurate linear registration and motion correction of brain images. NeuroImage, 17(2), 825–841. 10.1016/s1053-8119(02)91132-8

Jenkinson, M., Beckmann, C. F., Behrens, T. E., Woolrich, M. W., & Smith, S. M. (2012). FSL. NeuroImage, 62(2), 782–790. 10.1016/j.neuroimage.2011.09.015

Kaczmarzyk, J., McClure, P., Zulfikar, W., Rana, A., Rajaei, H., Richie-Halford, A., Bansal, S., Jarecka, D., Lee, J., & Ghosh, S. (2023, October). Neuronets/nobrainer: 1.1.1. 10.5281/zenodo.8417516

Kassambara, A. (2025, June). Ggpubr: ‘ggplot2’ Based Publication Ready Plots.

Keller, S. S., Ahrens, T., Mohammadi, S., Möddel, G., Kugel, H., Bernd Ringelstein, E., & Deppe, M. (2011). Microstructural and volumetric abnormalities of the putamen in juvenile myoclonic epilepsy. Epilepsia, 52(9), 1715–1724. 10.1111/j.1528-1167.2011.03117.x

Keller, S. S., Richardson, M. P., O’Muircheartaigh, J., Schoene-Bake, J.-C., Elger, C., & Weber, B. (2015). Morphometric MRI alterations and postoperative seizure control in refractory temporal lobe epilepsy: Morphometry and Outcome in Epilepsy. Human Brain Mapping, 36(5), 1637–1647. 10.1002/hbm.22722

Keller, S. S., Schoene-Bake, J.-C., Gerdes, J. S., Weber, B., & Deppe, M. (2012). Concomitant Fractional Anisotropy and Volumetric Abnormalities in Temporal Lobe Epilepsy: Cross-Sectional Evidence for Progressive Neurologic Injury. PLoS ONE, 7(10), e46791. 10.1371/journal.pone.0046791

Konell, H. G., Junior, L. O. M., Dos Santos, A. C., & Salmon, C. E. G. (2024). Assessment of U-Net in the segmentation of short tracts: Transferring to clinical MRI routine. Magnetic Resonance Imaging, S0730-725X(24)00158–9. 10.1016/j.mri.2024.05.009

Korbmacher, M., Westlye, L. T., & Maximov, I. I. (2024, June). FreeSurfer version-shuffling can boost brain age predictions. 10.1101/2024.06.14.599070

Laird, A. R. (2021). Large, open datasets for human connectomics research: Considerations for reproducible and responsible data use. NeuroImage, 244, 118579. 10.1016/j.neuroimage.2021.118579

Larivière, S., Rodríguez-Cruces, R., Royer, J., Caligiuri, M. E., Gambardella, A., Concha, L., Keller, S. S., Cendes, F., Yasuda, C., Bonilha, L., Gleichgerrcht, E., Focke, N. K., Domin, M., von Podewills, F., Langner, S., Rummel, C., Wiest, R., Martin, P., Kotikalapudi, R., . . . Bernhardt, B. C. (2020). Network-based atrophy modeling in the common epilepsies: A worldwide ENIGMA study. Science Advances. 10.1126/sciadv.abc6457

LeCun, Y., Bengio, Y., & Hinton, G. (2015). Deep learning. Nature, 521(7553), 436–444. 10.1038/nature14539

Leek, N. J., Neason, M., Kreilkamp, B. a. K., de Bezenac, C., Ziso, B., Elkommos, S., Das, K., Marson, A. G., & Keller, S. S. (2021). Thalamohippocampal atrophy in focal epilepsy of unknown cause at the time of diagnosis. European Journal of Neurology, 28(2), 367–376. 10.1111/ene.14565

Li, A., Mueller, K., & Ernst, T. (2004). Methods for efficient, high quality volume resampling in the frequency domain. IEEE Visualization 2004, 3–10. 10.1109/VISUAL.2004.70

Lie, I. A., Kerklingh, E., Wesnes, K., van Nederpelt, D. R., Brouwer, I., Torkildsen, Ø., Myhr, K.-M., Barkhof, F., Bø, L., & Vrenken, H. (2022). The effect of gadolinium-based contrast-agents on automated brain atrophy measurements by FreeSurfer in patients with multiple sclerosis. European Radiology, 32(5), 3576–3587. 10.1007/s00330-021-08405-8

Madan, C. R. (2022). Scan Once, Analyse Many: Using Large Open-Access Neuroimaging Datasets to Understand the Brain. Neuroinformatics, 20(1), 109–137. 10.1007/s12021-021-09519-6

May, A., & Gaser, C. (2006). Magnetic resonance-based morphometry: A window into structural plasticity of the brain. Current Opinion in Neurology, 19(4), 407. 10.1097/01.wco.0000236622.91495.21

Mayberg, M., Green, M., Vasserman, M., Raichman, D., Belenky, E., Wolf, M., Shrot, S., Kiryati, N., Konen, E., & Mayer, A. (2022). Anisotropic neural deblurring for MRI acceleration. International Journal of Computer Assisted Radiology and Surgery, 17(2), 315–327. 10.1007/s11548-021-02535-6

McKavanagh, A., Kreilkamp, B. A., Chen, Y., Denby, C., Bracewell, M., Das, K., De Bezenac, C., Marson, A. G., Taylor, P. N., & Keller, S. S. (2021). Altered Structural Brain Networks in Refractory and Nonrefractory Idiopathic Generalized Epilepsy. Brain Connectivity. 10.1089/brain.2021.0035

McKavanagh, A., Ridzuan-Allen, A., Kreilkamp, B. A. K., Chen, Y., Manjón, J. V., Coupé, P., Bracewell, M., Das, K., Taylor, P. N., Marson, A. G., & Keller, S. S. (2023). Midbrain structure volume, estimated myelin and functional connectivity in idiopathic generalised epilepsy. Epilepsy & Behavior, 140, 109084. 10.1016/j.yebeh.2023.109084

McKinley, R., Wepfer, R., Aschwanden, F., Grunder, L., Muri, R., Rummel, C., Verma, R., Weisstanner, C., Reyes, M., Salmen, A., Chan, A., Wagner, F., & Wiest, R. (2021). Simultaneous lesion and brain segmentation in multiple sclerosis using deep neural networks. Scientific Reports, 11(1), 1087. 10.1038/s41598-020-79925-4

Mugler III, J. P., & Brookeman, J. R. (1990). Three-dimensional magnetization-prepared rapid gradient-echo imaging (3D MP RAGE). Magnetic Resonance in Medicine, 15(1), 152–157. 10.1002/mrm.1910150117

Papanicolas, I., Woskie, L. R., & Jha, A. K. (2018). Health Care Spending in the United States and Other High-Income Countries. JAMA, 319(10), 1024–1039. 10.1001/jama.2018.1150

Pegg, E. J., McKavanagh, A., Bracewell, R. M., Chen, Y., Das, K., Denby, C., Kreilkamp, B. A. K., Laiou, P., Marson, A., Mohanraj, R., Taylor, J. R., & Keller, S. S. (2021). Functional network topology in drug resistant and well-controlled idiopathic generalized epilepsy: A resting state functional MRI study. Brain Communications, 3(3), fcab196. 10.1093/braincomms/fcab196

R Core Team. (2023). R: A Language and Environment for Statistical Computing.

Raghunathan, A., Xie, S. M., Yang, F., Duchi, J., & Liang, P. (2020, July). Understanding and Mitigating the Tradeoff Between Robustness and Accuracy. 10.48550/arXiv.2002.10716

Ratcliffe, C., Wandschneider, B., Baxendale, S., Thompson, P., Koepp, M. J., & Caciagli, L. (2020). Cognitive Function in Genetic Generalized Epilepsies: Insights From Neuropsychology and Neuroimaging. Frontiers in Neurology, 11, 144. 10.3389/fneur.2020.00144

Rebsamen, M., Capiglioni, M., Hoepner, R., Salmen, A., Wiest, R., Radojewski, P., & Rummel, C. (2023). Growing importance of brain morphometry analysis in the clinical routine: The hidden impact of MR sequence parameters. Journal of Neuroradiology = Journal De Neuroradiologie, S0150-9861(23)00198–. 10.1016/j.neurad.2023.04.003

Rebsamen, M., McKinley, R., Radojewski, P., Pistor, M., Friedli, C., Hoepner, R., Salmen, A., Chan, A., Reyes, M., Wagner, F., Wiest, R., & Rummel, C. (2022). Reliable brain morphometry from contrast-enhanced T1w-MRI in patients with multiple sclerosis. Human Brain Mapping, 44(3), 970–979. 10.1002/hbm.26117

Rebsamen, M., Rummel, C., Reyes, M., Wiest, R., & McKinley, R. (2020). Direct cortical thickness estimation using deep learning-based anatomy segmentation and cortex parcellation. Human Brain Mapping, 41(17), 4804–4814. 10.1002/hbm.25159

Revelle, W. (2023). Psych: Procedures for psychological, psychometric, and personality research. Rodriguez-Cruces, R., Royer, J., Larivière, S., Bassett, D. S., Caciagli, L., & Bernhardt, B. C. (2022). Multi-modal connectome biomarkers of cognitive and affective dysfunction in the common epilepsies. Network Neuroscience, 6(2), 320–338. 10.1162/netn_a_00237

Rorden, C. (2021). Surf Ice: A Simple Tool for Visualizing Connectome Networks, Tractography, and Statistical Maps.

Rorden, C., & Brett, M. (2000). Stereotaxic Display of Brain Lesions. Behavioural Neurology, 12(4), 191–200. 10.1155/2000/421719

Royer, J., Bernhardt, B. C., Larivière, S., Gleichgerrcht, E., Vorderwülbecke, B. J., Vulliémoz, S., & Bonilha, L. (2022). Epilepsy and brain network hubs. Epilepsia, n/a(n/a). 10.1111/epi.17171

RStudio Team. (2020). RStudio: Integrated development environment for R.

Rusak, F., Santa Cruz, R., Lebrat, L., Hlinka, O., Fripp, J., Smith, E., Fookes, C., Bradley, A. P., & Bourgeat, P. (2022). Quantifiable brain atrophy synthesis for benchmarking of cortical thickness estimation methods. Medical Image Analysis, 82, 102576. 10.1016/j.media.2022.102576

Samuel, G., Lucivero, F., & Lucassen, A. M. (2022). Sustainable biobanks: A case study for a green global bioethics. Global Bioethics, 33(1), 50–64. 10.1080/11287462.2021.1997428

Schaper, F. L. W. V. J., Nordberg, J., Cohen, A. L., Lin, C., Hsu, J., Horn, A., Ferguson, M. A., Siddiqi, S. H., Drew, W., Soussand, L., Winkler, A. M., Simó, M., Bruna, J., Rheims, S., Guenot, M., Bucci, M., Nummenmaa, L., Staals, J., Colon, A. J., . . . Fox, M. D. (2023). Mapping Lesion-Related Epilepsy to a Human Brain Network. JAMA neurology. 10.1001/jamaneurol.2023.1988

Scheffer, I. E., Berkovic, S., Capovilla, G., Connolly, M. B., French, J., Guilhoto, L., Hirsch, E., Jain, S., Mathern, G. W., Moshé, S. L., Nordli, D. R., Perucca, E., Tomson, T., Wiebe, S., Zhang, Y.-H., & Zuberi, S. M. (2017). ILAE classification of the epilepsies: Position paper of the ILAE Commission for Classification and Terminology. Epilepsia, 58(4), 512–521. 10.1111/epi.13709

Smith, S. (2002). Fast robust automated brain extraction. Human Brain Mapping, 17(3), 143–155. 10.1002/hbm.10062

Smith, S. M., & Nichols, T. E. (2009). Threshold-free cluster enhancement: Addressing problems of smoothing, threshold dependence and localisation in cluster inference. NeuroImage, 44(1), 83–98. 10.1016/j.neuroimage.2008.03.061

Spitzer, H., Ripart, M., Whitaker, K., D’Arco, F., Mankad, K., Chen, A. A., Napolitano, A., De Palma, L., De Benedictis, A., Foldes, S., Humphreys, Z., Zhang, K., Hu, W., Mo, J., Likeman, M., Davies, S., Güttler, C., Lenge, M., Cohen, N. T., . . . Wagstyl, K. (2022). Interpretable surface-based detection of focal cortical dysplasias: A Multi-centre Epilepsy Lesion Detection study. Brain: A Journal of Neurology, awac224. 10.1093/brain/awac224

The GIMP Development Team. (2019, June). GIMP.

The Scribus Team. (2023). Scribus: Open Source Desktop Publishing.

Tian, Q., Bilgic, B., Fan, Q., Ngamsombat, C., Zaretskaya, N., Fultz, N. E., Ohringer, N. A., Chaudhari, A. S., Hu, Y., Witzel, T., Setsompop, K., Polimeni, J. R., & Huang, S. Y. (2021). Improving in vivo human cerebral cortical surface reconstruction using data-driven super-resolution. Cerebral Cortex, 31(1), 463–482. 10.1093/cercor/bhaa237

Tor-Díez, C., Pham, C.-H., Meunier, H., Faisan, S., Bloch, I., Bednarek, N., Passat, N., & Rousseau, F. (2019). Evaluation of cortical segmentation pipelines on clinical neonatal MRI data. 2019 41st Annual International Conference of the IEEE Engineering in Medicine and Biology Society (EMBC), 6553–6556. 10.1109/EMBC.2019.8856795

Tustison, N. J., Avants, B. B., Cook, P. A., Song, G., Das, S., van Strien, N., Stone, J. R., & Gee, J. C. (2013). The ANTs cortical thickness processing pipeline. Medical Imaging 2013: Biomedical Applications in Molecular, Structural, and Functional Imaging, 8672, 126–129. 10.1117/12.2007128

Tustison, N. J., Avants, B. B., Cook, P. A., Zheng, Y., Egan, A., Yushkevich, P. A., & Gee, J. C. (2010). N4ITK: Improved N3 bias correction. IEEE transactions on medical imaging, 29(6), 1310–1320. 10.1109/TMI.2010.2046908

Tustison, N. J., Cook, P. A., Klein, A., Song, G., Das, S. R., Duda, J. T., Kandel, B. M., van Strien, N., Stone, J. R., Gee, J. C., & Avants, B. B. (2014). Large-scale evaluation of ANTs and FreeSurfer cortical thickness measurements. NeuroImage, 99, 166–179. 10.1016/j.neuroimage.2014.05.044

Van Horn, J. D., & Toga, A. W. (2014). Human Neuroimaging as a “Big Data” Science. Brain imaging and behavior, 8(2), 323–331. 10.1007/s11682-013-9255-y

Vidal-Jordana, A., Pareto, D., Sastre-Garriga, J., Auger, C., Ciampi, E., Montalban, X., & Rovira, A. (2017). Measurement of Cortical Thickness and Volume of Subcortical Structures in Multiple Sclerosis: Agreement between 2D Spin-Echo and 3D MPRAGE T1-Weighted Images. AJNR: American Journal of Neuroradiology, 38(2), 250–256. 10.3174/ajnr.A4999

Whelan, C. D., Altmann, A., Botía, J. A., Jahanshad, N., Hibar, D. P., Absil, J., Alhusaini, S., Alvim, M. K. M., Auvinen, P., Bartolini, E., Bergo, F. P. G., Bernardes, T., Blackmon, K., Braga, B., Caligiuri, M. E., Calvo, A., Carr, S. J., Chen, J., Chen, S., . . . Sisodiya, S. M. (2018). Structural brain abnormalities in the common epilepsies assessed in a worldwide ENIGMA study. Brain, 141(2), 391–408. 10.1093/brain/awx341

Wickham, H. (2016). Ggplot2: Elegant graphics for data analysis.

Wickham, H., Henry, L., Pedersen, T. L., Luciani, T. J., Decorde, M., & Lise, V. (2023). Svglite: An ‘SVG’ graphics device.

Wickham, H., & RStudio. (2023, January). Forcats: Tools for Working with Categorical Variables (Factors). Wickham, H., Vaughan, D., & Girlich, M. (2023). Tidyr: Tidy messy data.

Yao, A. D., Cheng, D. L., Pan, I., & Kitamura, F. (2020). Deep Learning in Neuroradiology: A Systematic Review of Current Algorithms and Approaches for the New Wave of Imaging Technology. Radiology: Artificial Intelligence, 2(2), e190026. 10.1148/ryai.2020190026

Yuan, J., Ran, X., Liu, K., Yao, C., Yao, Y., Wu, H., & Liu, Q. (2022). Machine learning applications on neuroimaging for diagnosis and prognosis of epilepsy: A review. Journal of Neuroscience Methods, 368, 109441. 10.1016/j.jneumeth.2021.109441

Zhao, C., Dewey, B. E., Pham, D. L., Calabresi, P. A., Reich, D. S., & Prince, J. L. (2021). SMORE: A Self-Supervised Anti-Aliasing and Super-Resolution Algorithm for MRI Using Deep Learning. IEEE Transactions on Medical Imaging, 40(3), 805–817. 10.1109/TMI.2020.3037187

Zubal, R., Velicky Buecheler, M., Sone, D., Postma, T., De Tisi, J., Caciagli, L., Winston, G. P., Sidhu, M. K., Long, L., Xiao, B., Mcevoy, A. W., Miserocchi, A., Vos, S. B., Baumann, C. R., Duncan, J. S., Koepp, M. J., & Galovic, M. (2025). Brain Hypertrophy in Patients With Mesial Temporal Lobe Epilepsy With Hippocampal Sclerosis and Its Clinical Correlates. Neurology, 104(2), e210182. 10.1212/WNL.0000000000210182

