## Supplementary figures and images for "Machine Learning-Based Reconstruction of 2D MRI for Quantitative Morphometry in Epilepsy"

### Figure a1

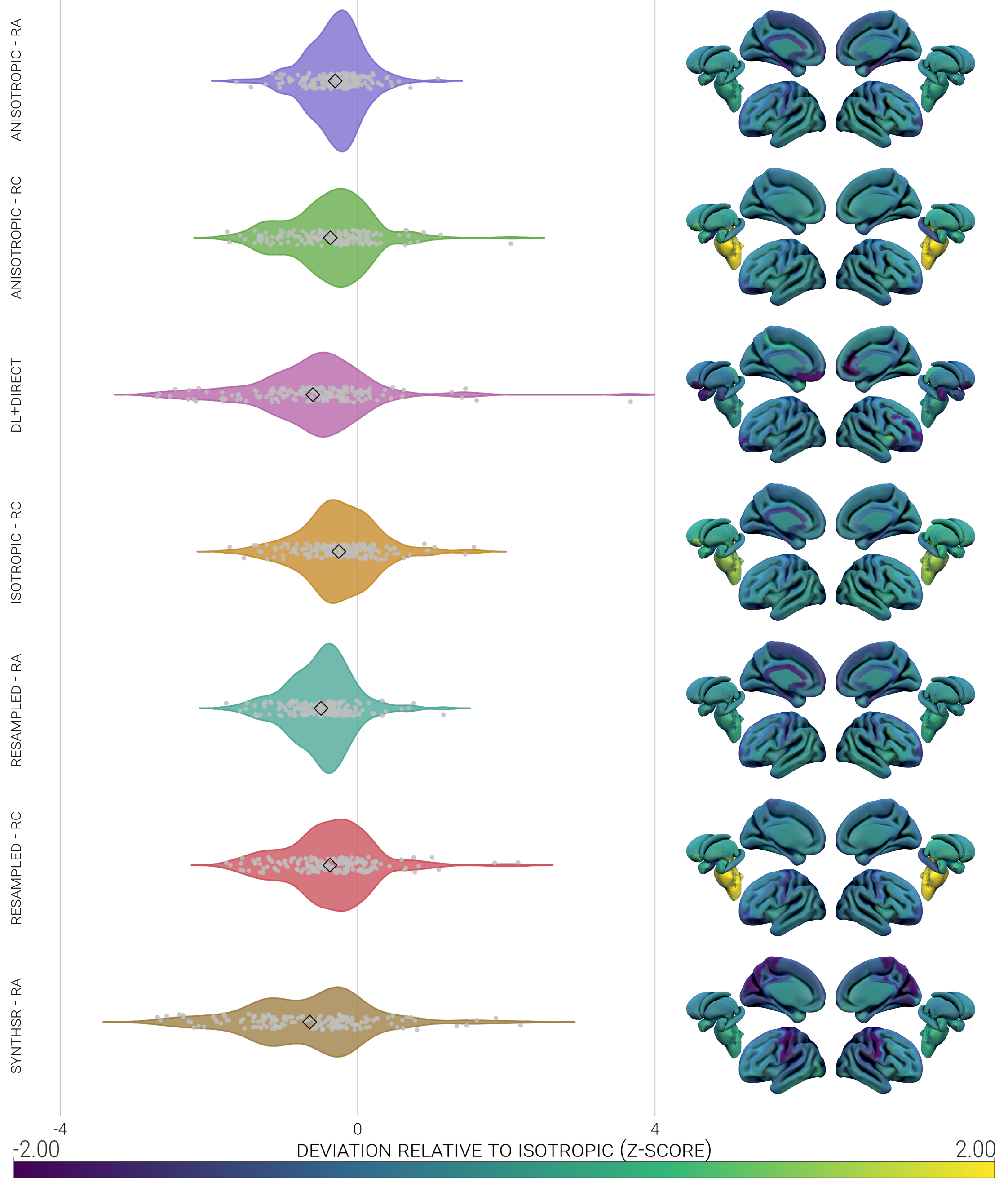

### Figure a2

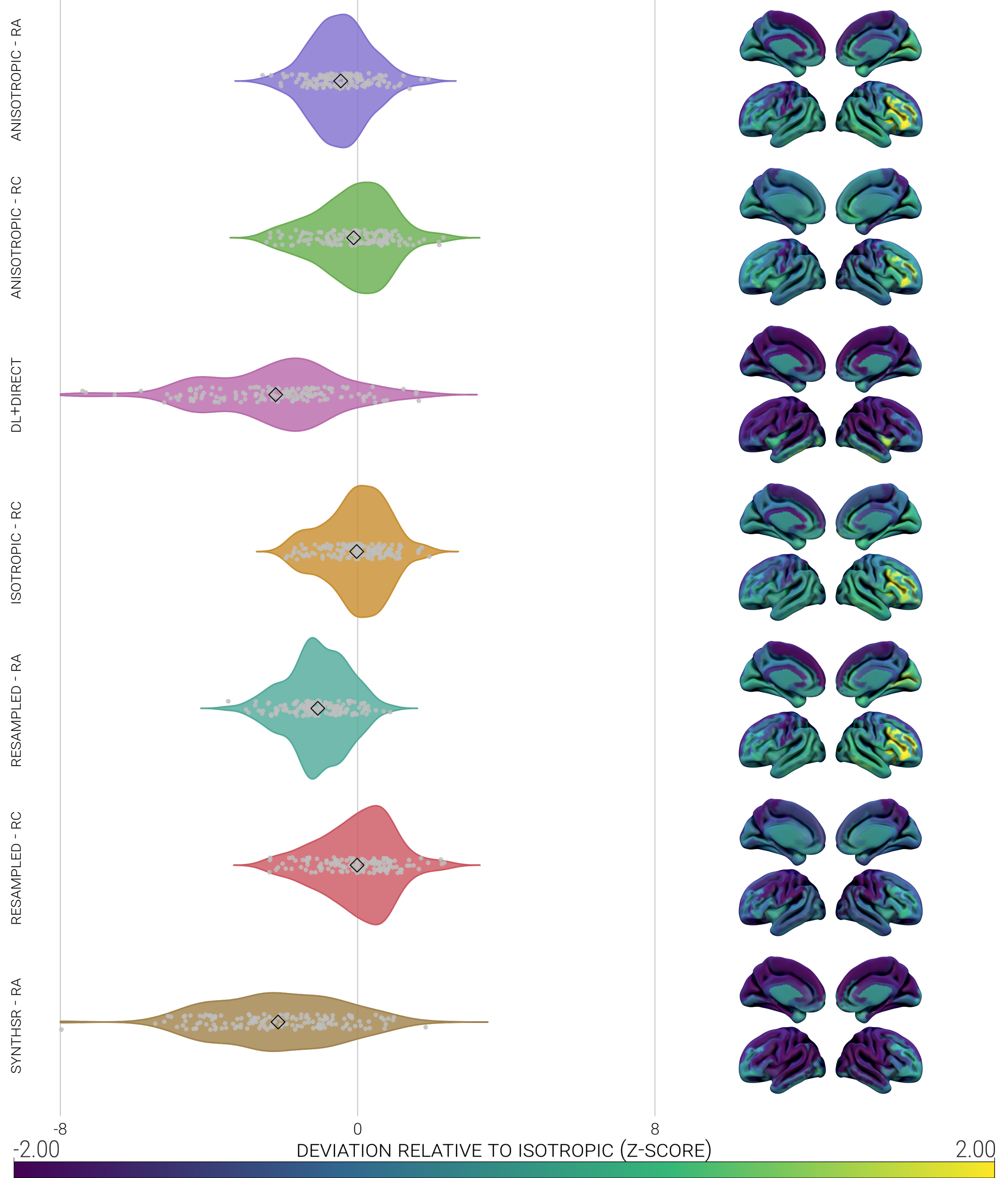

### Figure a3

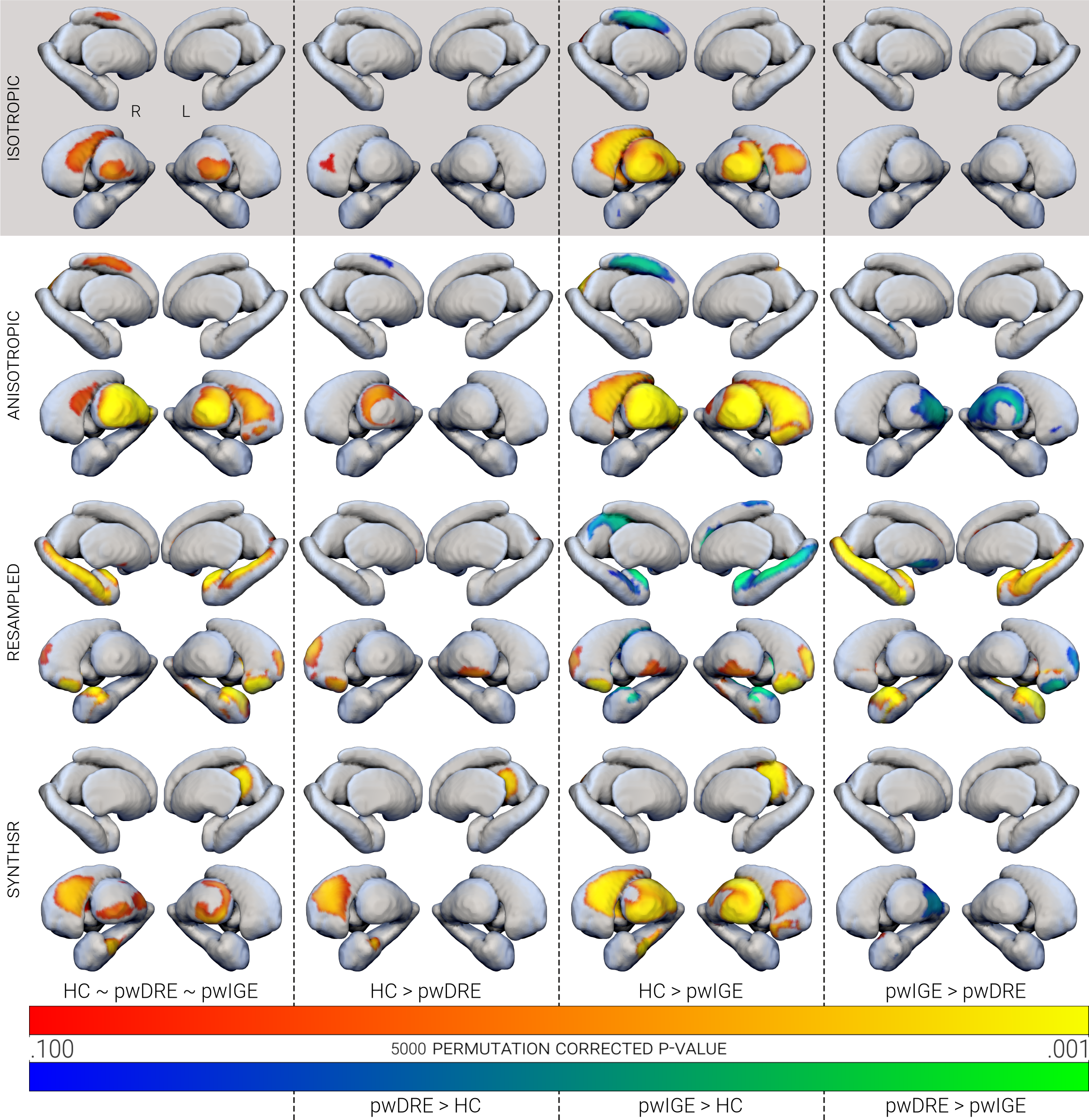
